# Evolution of child acute malnutrition during war in the Gaza Strip, 2023-2024: retrospective estimates and scenario-based projections

**DOI:** 10.1101/2024.12.10.24318783

**Authors:** Francesco Checchi, Zeina Jamaluddine

**Affiliations:** Faculty of Epidemiology and Population Health, London School of Hygiene and Tropical Medicine, London, UK

**Keywords:** Gaza, occupied Palestinian territories, war, conflict, insecurity, calorie, nutrition, wasting, mathematical model

## Abstract

**Background:** Nutritional status has been compromised by ongoing war and restrictions on food deliveries in the Gaza Strip. We developed a mathematical model that outputs retrospective estimates and scenario-based projections of acute malnutrition prevalence among children given caloric intake and other factors. We present here the model and its application to the crisis in Gaza.

**Methods:** We extended an existing mechanistic model for weight change as a function of energy balance, calibrating it to represent variability in growth curves observed in pre-war Gaza. We simulated open cohorts of children exposed to time-varying caloric intake, infant exclusive breastfeeding prevalence, incidence of infectious disease and coverage of malnutrition treatment, while allowing for adult caloric sacrifice to supplement child intake in times of food scarcity.

**Results and conclusions:** The model accurately replicates growth standards, pre-war growth patterns and expected parameter dependencies. It suggests that a considerable increase in acute malnutrition occurred in northern Gaza during early 2024. Projections for late 2024 include a serious nutritional emergency if relatively pessimistic assumptions are made about food availability. The model may hold considerable promise for informing decisions in humanitarian response but requires further validation and development.

## Introduction

Since October 2023, the Gaza Strip has endured intense military operations and severe restrictions on circulation of people and goods. During the first seven months of the war (October 2023 to April 2024), reduced food deliveries caused highly prevalent food insecurity and triggered warnings of impending famine [1, 2]. There were consistent media and civil society reports of food scarcity, particularly among people who had remained in the two northern governorates (Gaza City, North Gaza), where food deliveries had been scarcest, and instances of fatal starvation were recorded [3]. From April 2024 onwards, an apparent improvement in food security coinciding with re-opened border crossings was noted [4], but objective data were obscured by Israel’s takeover of customs points from which the United Nations had previously tracked the content of incoming truckloads [5].

Acute malnutrition is a short-term consequence of dietary insufficiency and, in crises, disproportionately affects children and pregnant or lactating women [6]. Its population burden is typically measured by cross-sectional surveys in which children aged 6 to 59 months old (mo) are classified as severely or moderately malnourished based on their weight-for-height Z-score (WHZ) and/or the presence of bilateral pitting oedema [7, 8]. In Gaza, these surveys have been hampered by insecurity; humanitarian actors have instead relied on rapid community-based screening based on middle-upper-arm circumference (MUAC), an anthropometric index that, while predictive of the risk of death [9], does not correlate well with WHZ [10]. These screenings have, to our knowledge, relied on convenience rather than population-representative samples.

In a separate publication, we analysed multiple sources of food to estimate caloric availability over time in both northern Gaza and the south-central governorates [11]. Here, we introduce a mathematical modelling framework to estimate acute malnutrition prevalence based on caloric availability, whilst accounting for protective or risk factors including malnutrition treatment, incidence of common infectious disease, reduced breastfeeding and disproportionate sharing of food by adults with their children (hereafter ‘caloric sacrifice’), a plausible coping mechanism. We apply the model to the Gaza context and reconstruct the evolution of acute malnutrition prevalence, while also forward-projecting it under alternative scenarios.

## Methods

### Study population and period

We stratified Gaza into a northern (North Gaza, Gaza City governorates) and south-central region (Deir al Balah, Khan Younis, Rafah). The northern region contained approximately 1,200,000 people pre-war [12] but only about 250,000 by March 2024 [13]. We sourced Gaza’s age-sex distribution and proportion of pregnant and lactating women from UN projections of the 2017 census [14] and the distribution and mean number of household members from the census itself [15].

Our retrospective analysis period spans 7 October 2023 to 6 May 2024, when the Israeli Defence Force (IDF)’s takeover of the southern border crossings meant the UN could no longer collect comprehensive food trucking data [5]. We present scenario-based forward projections for the period from 7 May to 31 December 2024.

### Overall model description

We simulate an open cohort of children 0 to 59mo with daily time steps. Each child is attributed a random age, sex and height and weight growth percentiles at birth that determine their anthropometric trajectory through age and time under non-crisis conditions. These percentiles are based on the estimated distribution of growth curves for pre-war Gaza (see below), which are assumed to already reflect exposure to pre-crisis levels of different risk factors and used to estimate pre-war caloric intake. The cohort is aged up to the crisis’ start date and is then progressed through the retrospective estimation period by exposing it to the estimated caloric availability per day during the crisis, adjusted for the extent of adults’ assumed caloric sacrifice, as well as time-varying factors including *excess* (i.e. crisis minus baseline) burden of acute respiratory infection and acute watery diarrhoea, excess prevalence of non-exclusive breastfeeding during infancy, and the coverage of severe and moderate acute malnutrition (SAM, MAM) treatment. The model then exposes the cohort to one or more forward-projection scenarios in which the values of the above variables are arbitrarily set to explore possible crisis trajectories.

Children who age out of the cohort are replaced by newborns with the same sex, breastfeeding exclusivity and height-weight percentiles. Mortality is omitted and population assumed constant. We adapt a published mechanistic model [16] to predict children’s weight change over each time step as a function of caloric intake and resulting energy balance. We present model components and inputs below. Data and code for implementation in R [17] are publicly available on https://github.com/francescochecchi/nut_cal_model_gaza.

### Pre-war growth curve estimation

Before the war, the United Nations Relief and Works Agency for Palestine Refugees in the Near East (UNRWA) offered free growth monitoring visits to Gazan children up to age 5y. We sourced from UNRWA 2,704,336 (1,307,754 female or 48.8%) individual weight and height observations collected longitudinally between 2019 and 2023 (a median 10 observations per child), reduced to 2,697,477 (99.7%) after removing children with <3 observations and implausible values (weight-for-height, weight-for-age and/or height-for-age <> 5 Z-scores as computed using the R `anthro` package developed to accompany the World Health Organization’s 2006 growth standards [18]). As shown in the Supplementary Material (Figure S6), observations were mostly spaced three months apart and decreased after age 18mo.

We applied to the above data the WHO’s method for constructing a pre-war distribution of sex-specific weight-for-age (WAZ) and height-for-age (HAZ) growth curves (Figure S7, Figure S8), which relies on a generalised additive model for location, size and shape (GAMLSS, implemented through the `gamlss` package [19]) with a Box-Cox-power-exponential distribution [18]. From each model fit we extracted the predicted 0-99% percentiles of weight and height at any age.

We assumed that children would remain at the same percentile through life if pre-war conditions persisted. In reality, children exhibit individual heterogeneity in their trajectory, but our simplifying assumption should be appropriate as long as it accurately replicates observed population variability (see Results): this should occur if heterogeneity is random. Accordingly, we allocated an equal fraction of the cohort to each weight percentile. We also attributed a random height percentile, accounting for the observed correlation of height with weight curves (Figure S9). We did this by first fitting (in package `mgcv`[20]) a generalised additive negative-binomial model to the count of children within each mean weight-height percentile combination, featuring a tensor smoothing product of each child’s mean weight and height percentile as observed over their longitudinal observations, adjusted for sex and with the log of the number of children in each mean weight percentile as an offset (Figure S10). We used the model-fitted counts of height percentile within each weight percentile as a probability distribution from which to sample when attributing a random birth height percentile to each child in the cohort, conditional on weight percentile. We also tried sampling from the empirical weight-height correlation matrix, but this resulted in cohorts with height-for-age variance well below that observed.

### Weight sub-model

Hall et al. [16] have developed a mechanistic model that predicts weight change over time *t* as the sum of change in fat (FM) and fat-free (lean) mass (FFM), given the balance of energy (caloric) intake and energy expenditure (*I*_*t*_ − *E*_*t*_). Briefly, *E*_*t*_ sums requirements due to baseline metabolism, food digestion, physical activity and growth, which vary by sex and age based on set constants. FM and FFM are burnt if intake is insufficient and accrete if intake surpasses expenditure; both processes entail energy costs and gains. Partitioning of energy between FM and FFM is age-dependent and a function of their relative availability. The model, developed to support weight management programmes, has been validated in older children and adults, but to our knowledge not applied widely among young children (see Discussion). We verified that it satisfactorily predicted the expected WHO standards by setting birth weight at the 0 WAZ level as per WHO standards and FM and FFM at birth based on Fomon et al.’s expected median values [21], and running the model to age 60mo (Figure S11). When applying the model to Gaza, we set birth weight percentiles as outlined above, and apportioned birth weight into FM and FFM using the sex-specific regression equations provided by Eriksson et al. [22].

Unless a caloric deficit/surplus is specified, the weight model outputs growth under energy balance, i.e. its predictions default to the population median no matter which birth weight a child starts out with (Figure S13). This masks the expected variability of WAZ and compromises the model’s application for SAM and GAM prevalence estimation, which relies on accurately estimating not just the central tendency but also the distribution of WAZ, HAZ and thus WHZ, so that SAM and GAM cases, who exist at the negative end of the curve, may be modelled accurately [23]. To correctly replicate the weight trajectories corresponding to different growth percentiles, we introduced a sex (*s*)-, age (*a*)- and weight percentile (*w*)-dependent calibration factor *m*_*s,a,w*_ defined by a two-parameter cumulative log-normal function, which seemed suitable for the pattern evident in growth curves, namely rapid weight gain in infancy followed by lower weight gain velocity:

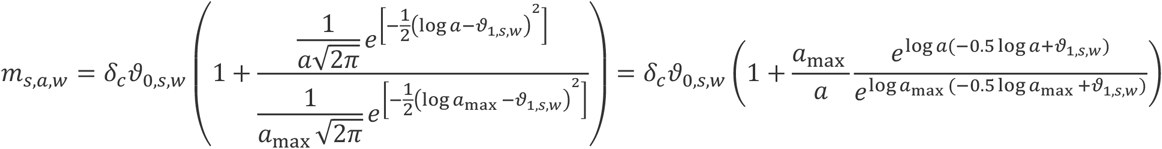

where ϑ_0,*s,w*_ and ϑ_1,*s,w*_ are unknown parameters and 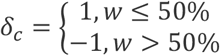.

When predicting weight change, we calibrated daily energy expenditure 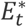 as *E*_*t*_(1 + *m*_*s,a,w*_), such that children at low weight percentiles were modelled as less energy-efficient, and vice versa. We estimated ϑ_0,*s,w*_ and ϑ_1,*s,w*_ by two-dimensional grid search, minimising the root sum of squares of model-predicted versus GAMLSS-fitted weight-for-age curves at each percentile, under each pair of candidate parameter values. Maximum-likelihood values, shown in Figure S14, were then associated with each weight percentile in further model application. As shown in Figure 1 for representative percentiles, the model thusly calibrated closely replicated GAMLSS-fitted growth percentiles.

**Figure 1.**
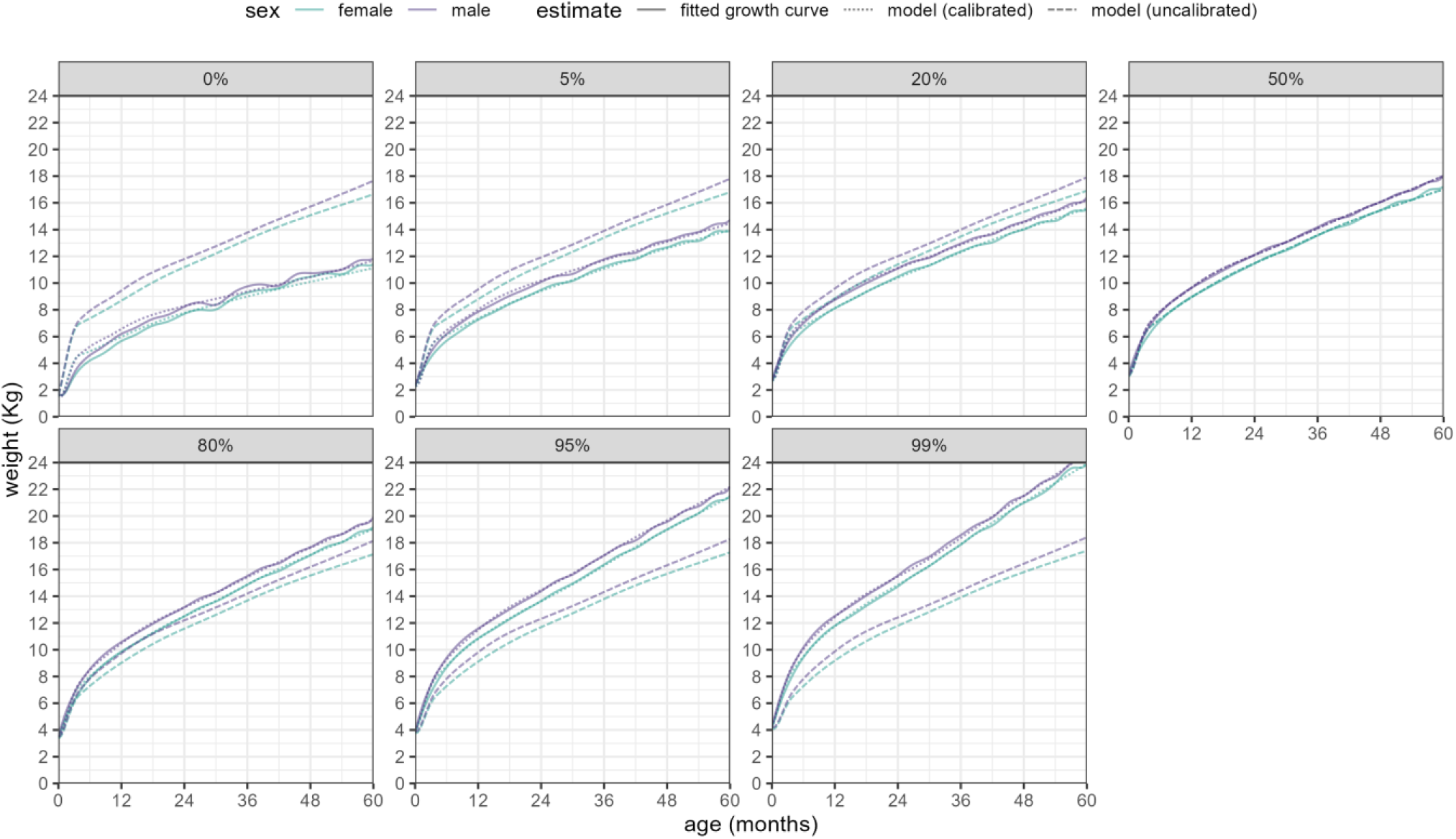
Fit of calibrated versus uncalibrated weight model predictions, by age and sex, compared to fitted growth curves, for a selection of weight percentiles.

### Other model components

#### Adult caloric sacrifice

We assumed that, in a situation of scarcity, adults (≥20yo) within each household would sacrifice a proportion of their available food to feed children, as shown in Gaza [1] and elsewhere [24, 25]. To work out the extent to which this sacrifice would increase children’s intake (as a multiplier adjustment factor *f*), we simulated 1000 households with size randomly sampled from the distribution of pre-war household size in Gaza, comprising of at least one adult and with remaining members’ age and sex allocated randomly based on Gaza’s age-sex distribution, with a Poisson-distributed random number of pregnant and lactating women, based on their population prevalence. We explored how *f* would vary as a function of (i) the proportion of their recommended intake that adults would be willing to sacrifice, (ii) the proportion of children’s recommended intake that adults would try to safeguard and (iii) the mean available per-capita Kcal/day. We assumed that food would be distributed equally among households based on their requirement (i.e. the sum of each member’s recommended intake), and that within households food would flow from adults to children until either the former’s sacrifice limit or the latter’s intake target were reached. As shown in Figure 2, a substantial adjustment to children’s intake would only occur when food is scarce yet enough that adults can spare some; below ≈600 Kcal/day, adults themselves are unlikely to afford sharing food, as this is already about one fourth of their recommended intake.

**Figure 2.**
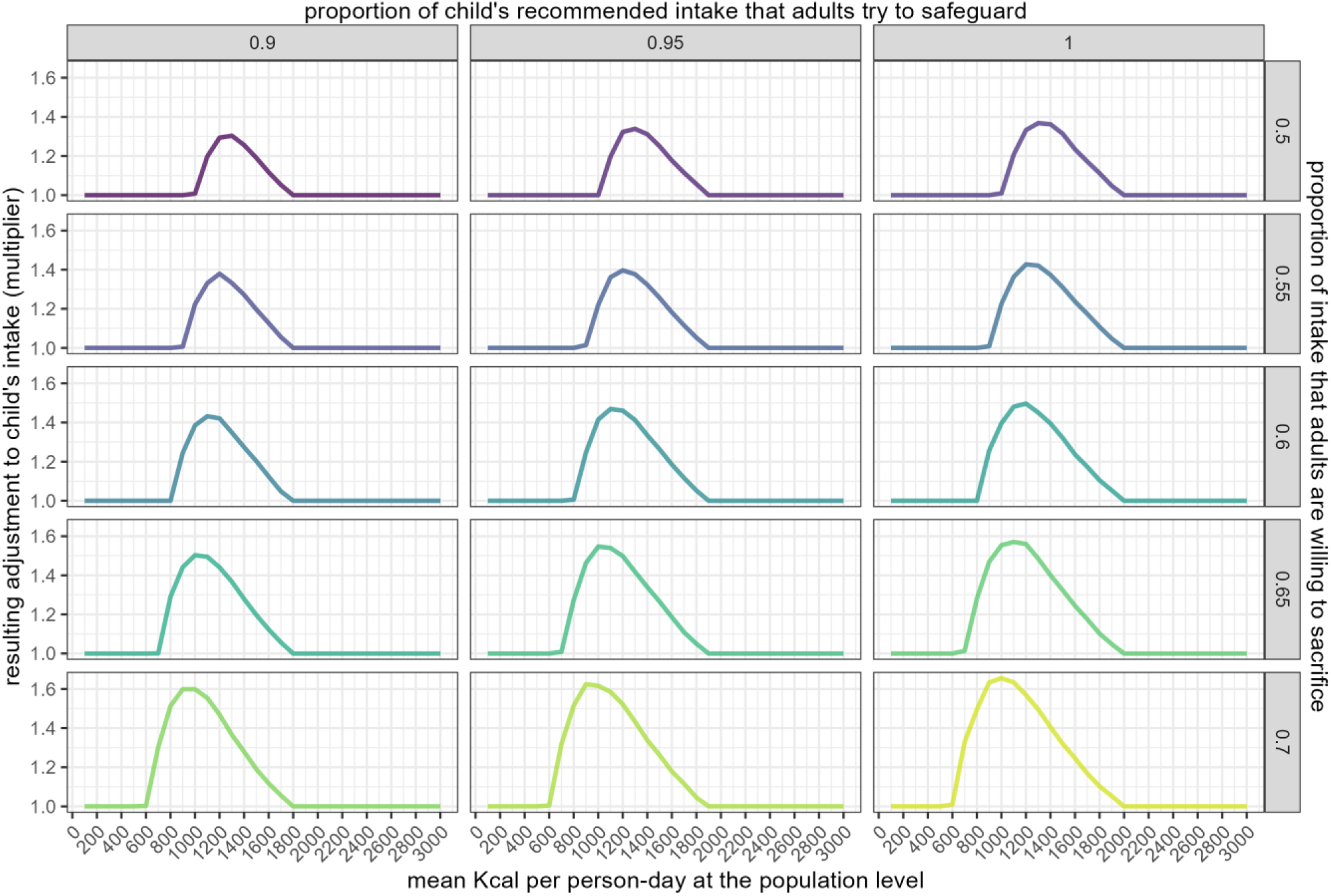
Adjustment (multiplier) to child’s daily caloric intake resulting from different levels of mean caloric availability at the population level (x-axis), maximum proportion of adult intake sacrificed (rows) and proportion of children’s recommended intake that adults will try to safeguard (columns).

#### Effect of infectious disease

We modelled the effect of common endemic infectious diseases as a reduction in daily caloric intake applied over each disease episode’s duration. After a non-systematic key word search of Medline, we identified five cohort studies of children with acute watery diarrhoea [26–29] and one of children with ARI [27], both conditions broadly defined according to WHO syndromic definitions, in which intake was also measured; we averaged these studies’ estimates of the ratio of intake when ill to intake when healthy.

To simulate disease episodes, we adapted Schmidt et al.’s method [30], whereby children are attributed a random annual incidence drawn from a gamma distribution whose shape parameter is the average local incidence and whose scale parameter captures expected heterogeneity. Children are also attributed a random episode duration (in days), itself drawn from a gamma distribution but adjusted (i) for observed positive correlation of duration and incidence based on a linear regression coefficient and (ii) for individual variability, based on a random normal deviate. We sourced the scale parameter of incidence, the shape and scale parameters of duration and the regression coefficient of duration by incidence from studies reviewed by the same authors [30].

Because incidence is rarely if ever measured in crisis settings, we worked out its correspondence with more survey-amenable indicators (point prevalence or period prevalence based on caregiver recall of symptoms over the previous two weeks) at various levels within a realistic range (Figure S15) by running a simulation of 1000 children over two years, with a burn-in period of one year to achieve steady state, in which for different incidence levels we computed corresponding prevalences.

Within the main model cohort, we set a daily point prevalence of ARI and diarrhoea, and worked out how many children would newly fall ill to make up this prevalence, after subtracting prevalent cases. We attributed new cases based on individual probabilities equal to zero if children were already ill (this prevented episode overlap) and the ratio of the child’s incidence to the mean incidence otherwise (to reproduce heterogeneity). As mentioned above, prevalence was specified as an excess level (crisis – baseline). The model does not currently accommodate a reduction from pre-crisis levels.

#### Effect of incomplete breastfeeding

We represented the effect of non-exclusive breastfeeding as an increase in the energy requirement of infants below 6mo, reflecting the lower caloric efficient of formula versus breastmilk; multipliers were taken from the Food and Agriculture Organisation [31] (Figure S16). As above, the excess prevalence of non-exclusive breastfeeding was specified.

#### Acute malnutrition treatment

While various protocols for SAM and MAM management exist, we simulated the simplified outpatient version with weekly follow-up common in acute emergency responses [32], whereby SAM cases receive 1000 Kcal/day (reduced to 500 if they improve to and stay at MAM over two consecutive weeks) while MAM cases receive 500 Kcal/day; treatment ends for both SAM and MAM if cases maintain a WHZ≥-2 over two consecutive weeks. We assumed that children would experience a delay before accessing treatment, sampled from a lognormal distribution with mean 15d and standard deviation equal to half the mean. We also assumed that children on treatment would continue to consume non-therapeutic food. We ignored additional therapeutic food that may be given in inpatient settings (e.g. specialised milk), as most SAM cases are usually treated on an outpatient basis; we also omitted MUAC and oedema (an infrequent and geographically clustered presentation of acute malnutrition [33]) as alternative admission and discharge criteria, since the model cannot estimate these anthropometric indices. Whenever the actual number of children admitted to treatment was known, we calculated the daily mean number of admissions given the cohort size relative to the total population, and attributed these among as-yet untreated SAM/MAM cases in the cohort based on a random Poisson process. The model also allows for treatment coverage (proportion) as an input, obviating the need for population data; this option is implemented as a random binomial probability. For simplicity we assumed 100% cure and no defaulting.

### Crisis-specific data sources

#### Recommended and actual caloric intake

Recommended intake by age-sex, with additions for pregnant and lactating women, was sourced from WHO guidelines [34]. Hall et al. [16] however offer an analytic solution for the recommended intake by age-sex, given an expected age- and sex-dependent FM to FFM ratio among healthy children. We applied this solution to compute recommended intakes among children 0-10yo (Figure S12) and used them in lieu of WHO’s. Among girls and boys aged 0 to 4yo, a mean 1117 and 1284 Kcal-day were predicted by Hall et al. compared to 1250 and 1320 recommended by WHO, with corresponding figures among 5 to 9yo’s being 1629 and 1725 versus 1730 and 1980.

We estimated pre-war actual intake per capita by doing a grid search over a range of candidate values of the ratio of actual to recommended mean caloric intake and finding the value of this ratio (and thus the actual intake) that minimised the root mean square error (RMSE) of weight model predictions, compared to the median WAZ growth curve. While the estimated actual mean intake was only 2% less than the recommended value (Figure S17), this small adjustment graphically improved model fit to the median pre-war growth curves (Figure S18). Intake during the war is estimated in a separate paper [11] and introduced here as an empirical distribution of 1000 simulation run outputs per day for both northern and south-central Gaza, from which we sample over the retrospective analysis period.

#### Other factors

We sourced the number of MAM and SAM treatment admissions from a Nutrition Cluster dashboard [35]: these are available from January 2024, prior to which we assume that no MAM/SAM treatment was occurring. ARI and diarrhoea two-week period prevalences were estimated by the MICS before the war [36]; during the war, rapid surveys [37–39] also estimated this indicator intermittently [40] but with a simpler questionnaire compared to MICS, so we crudely assumed that only half of cases identified were indeed ARI and acute diarrhoea (i.e. 50% positive and 100% negative predictive values). Further disease assumptions are in Table S3 and Table S4. The MICS estimated a 58% prevalence of complete or partial formula feeding among infants <6mo, which we assumed to have risen to 70% by 1 February 2024 in line with observed deteriorations in exclusive breastfeeding across crises in the Middle East [41–44]. We assumed that adults would seek to safeguard 100% of children’s caloric requirement and sacrifice up to 80% of theirs on any given day. Lastly, we sourced population estimates by region from the Palestinian Central Bureau of Statistics [45], the Food Security Cluster [46], the United Nations Office for Coordination of Humanitarian Affairs, UNOCHA [47] and UNRWA [48], and interpolated these linearly to fill in periods of missingness.

### Scenario assumptions

Table 1 details values and assumptions for a central (most plausible), reasonable-best and reasonable-worst scenarios. For simplicity we assumed that during the projection period (May to December 2024) only intake (Figure S19) and malnutrition treatment would vary by scenario, with ARI and diarrhoea prevalence as observed in the previous season (Table S3, Table S4) and all other factors at their April 2024 level.

**Table 1.** Scenarios for projection, by factor, sub-period and region of Gaza.

| Factor | Scenario | Sub-period (2024) | North |  | South-central |  |
| --- | --- | --- | --- | --- | --- | --- |
|  |  |  | Value | Justification | Value | Justification |
| Caloric intake | Reasonable best | 7 May to 30 Sep | unchanged relative to April | assumed; as April levels were higher than pre-war, this is considered a ceiling | Update of Checchi et al. [11] analysis, with trucked-in Kcal divided by (1 – underreporting) | underreporting = 44% (see text) |
|  |  | 1 Oct to 31 Dec | 40% decrease relative to April | considerably reduced supply due to renewed IDF offensive | 20% decrease relative to June-September | moderately reduced supply from southern crossings |
|  | Central (plausible) | 7 May to 30 Sep | 10% decrease relative to April | assumed | as for reasonable-best scenario | underreporting = 33% (see text) |
|  |  | 1 Oct to 31 Dec | 50% decrease relative to April | mid-point of best and worst scenarios | 30% decrease relative to June-September | mid-point of best and worst scenarios |
|  | Reasonable worst | 7 May to 30 Sep | 20% decrease relative to April | assumed | as for reasonable-best scenario | underreporting = 22% (see text) |
|  |  | 1 Oct to 31 Dec | 60% decrease relative to April | severely reduced supply due to renewed IDF offensive | 40% decrease relative to June-September | severely reduced supply from southern crossings |
| SAM and MAM treatment admissions | Reasonable best | 7 May to 30 Sep | (known) |  | (known) |  |
|  |  | 1 Oct to 31 Dec | 25% decrease in accessible treatment points relative to September | increased insecurity in northern Gaza | same number of accessible treatment points as in September | stable humanitarian operations |
|  | Central (plausible) | 7 May to 30 Sep | (known) |  | (known) |  |
|  |  | 1 Oct to 31 Dec | 50% decrease in accessible treatment points relative to September | increased insecurity in northern Gaza | 25% decrease in accessible treatment points relative to September | moderately reduced supply from southern crossings |
|  | Reasonable worst | 7 May to 30 Sep | (known) |  | (known) |  |
|  |  | 1 Oct to 31 Dec | 75% decrease in accessible treatment points relative to September | increased insecurity in northern Gaza | 50% decrease in accessible treatment points relative to September | considerably reduced supply from southern crossings |

We had actual MAM/SAM treatment admissions up to 30 September 2024 [35]. Thereafter, we crudely calculated total treatment capacity as the number of functional outpatient and inpatient MAM/SAM treatment points [35] times up to 5 MAM and 5 SAM admissions/day, reducing this total by a certain percentage for each scenario.

From 7 May onwards UNRWA was unable to comprehensively monitor food truckloads into Gaza, particularly those from the commercial sector, and appeared to capture only truckloads into south-central governorates: accordingly, for the south-central region we updated our published caloric availability estimates up to 30 September, but upward-adjusted trucked-in amounts from 7 May by assuming underreporting fractions corresponding in the reasonable-best scenario to the proportion of food coming from commercial sources during May-September 2024, namely 44% as reported by UNOCHA [49], and in the reasonable-worst scenario to half that level, closer to the proportion of commercial deliveries *before* May. For northern Gaza, we assumed a level of caloric intake relative to April 2024. From early October, Israel intensified attacks on northern Gaza, halting civilian supplies [50]: this suggested a qualitatively different trajectory for this sub-period and region (Table 1).

### Ethics

The study was approved by the Ethics Committee of the London School of Hygiene and Tropical Medicine (ref. 29926).

## Results

### Model testing

Table 2 and Figure S20 compare the model’s simulated pre-war distributions of HAZ, WAZ and WHZ to those obtained from the UNRWA growth monitoring dataset and the 2019-2020 MICS, indicating reasonable concordance, albeit with a lower modelled-than-observed variance. Figure S21 compares the model to a pre-war survey done among children <2yo by Albelbeisi et al. [51].

**Table 2.** Model-predicted versus observed (growth monitoring) pre-war anthropometry, by index and source.

| index | source† | mean | SD | % < -2SD (GAM) | % < -3SD (SAM) | % missing / implausible |
| --- | --- | --- | --- | --- | --- | --- |
| height-for-age Z-score | model | -0.31 | 0.95 | 3.9% | 0.8% | 0.0% |
|  | observed | -0.29 | 1.16 | 5.3% | 1.3% | 0.1% |
|  | MICS | -0.5 | not provided | 8.6% | 2.4% | not provided |
| weight-for-age Z-score | model | -0.19 | 0.98 | 3.8% | 1.1% | 0.0% |
|  | observed | -0.16 | 1.04 | 4.2% | 1.1% | 0.1% |
|  | MICS | 0.0 | not provided | 2.1% | 0.3% | not provided |
| weight-for-height Z-score | model | -0.04 | 0.94 | 2.5% | 1.0% | 0.1% |
|  | observed | 0.01 | 1.12 | 3.5% | 0.8% | 0.3% |
|  | MICS | 0.6 | not provided | 0.8% | 0.4% | not provided |

† Model: 20,000 simulated children. Observed: 260,895 growth monitoring observations during 2019-2023 (one per child, selected randomly). MICS (Multiple Indicator Cluster Survey): 2534 children measured cross-sectionally in 2019.

As shown in Figure 3A, the model exhibits the expected behaviour as hypothetical scenarios progress from caloric requirements being exactly met (stable GAM prevalence) to unmitigated caloric insufficiency (steep increase), insufficiency with adult sacrifice, excess infectious disease and finally partial treatment coverage. Figure 3B illustrates the strong influence of adult caloric sacrifice in model dynamics (see Discussion).

**Figure 3.**
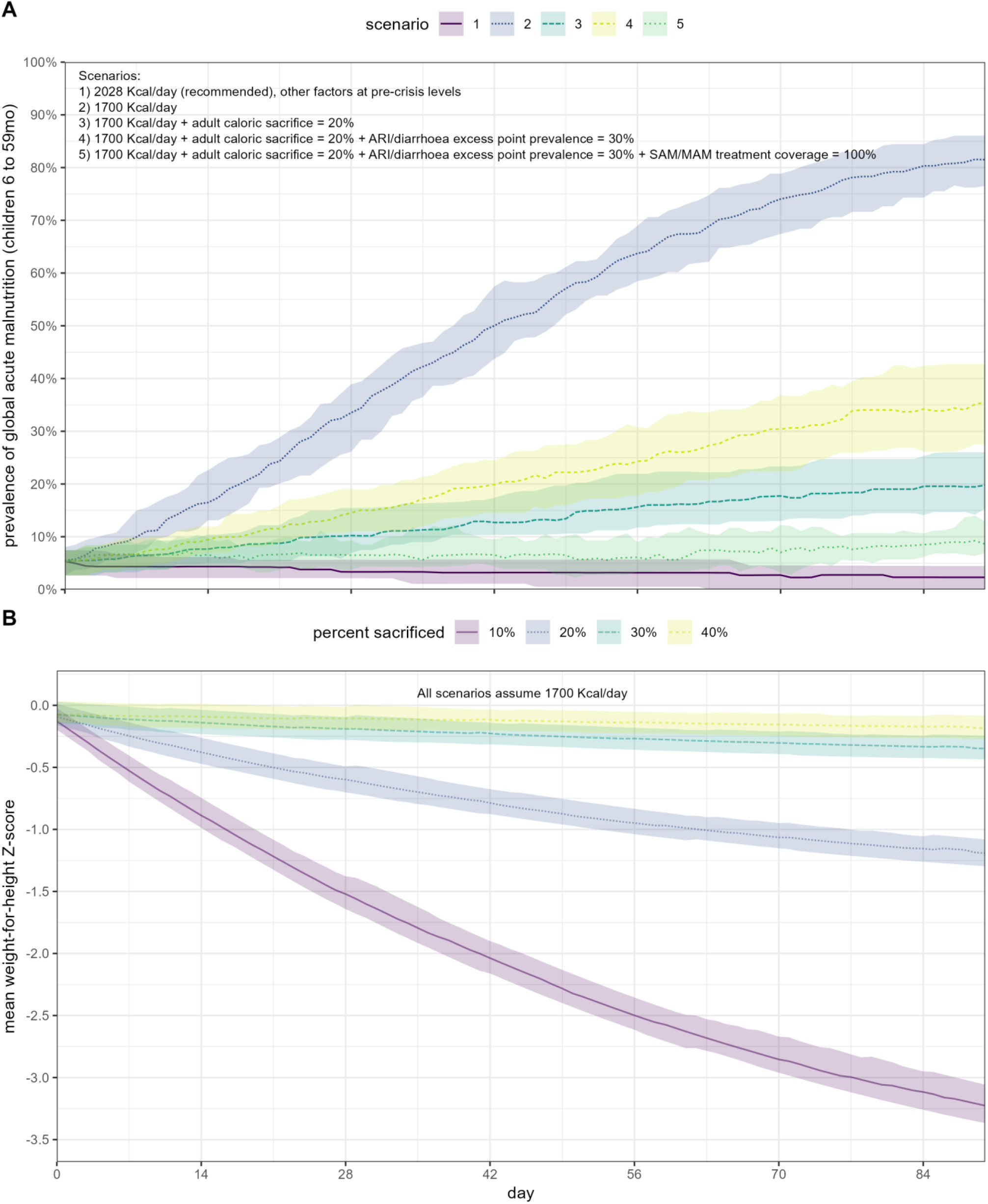
A: Modelled evolution of GAM prevalence under different scenarios. B: modelled evolution of mean WHZ under different values of adult caloric sacrifice, assuming mean intake 1700 Kcal/day and pre-crisis levels of all other factors. All scenarios comprise 20 simulated cohorts (runs) of 100 children and assume the same demographic characteristics as pre-war Gaza. Shaded areas denote 95% percentile intervals.

### Crisis estimates

As shown in Figure S22, the model generally yielded wide uncertainty intervals, especially for the retrospective period, during which uncertainty in food intake is propagated from our prior analysis. For easier readability, Figure 4 only reports point estimates. We estimated that acute malnutrition peaked around March 2024 in northern Gaza, with GAM prevalence considerably increased from its ≈4% pre-war modelled level (Table 2). Under the reasonable worst-case scenario, we projected that GAM and SAM prevalence would reach serious levels in both regions by December 2024, with a relatively higher increase in northern Gaza.

**Figure 4.**
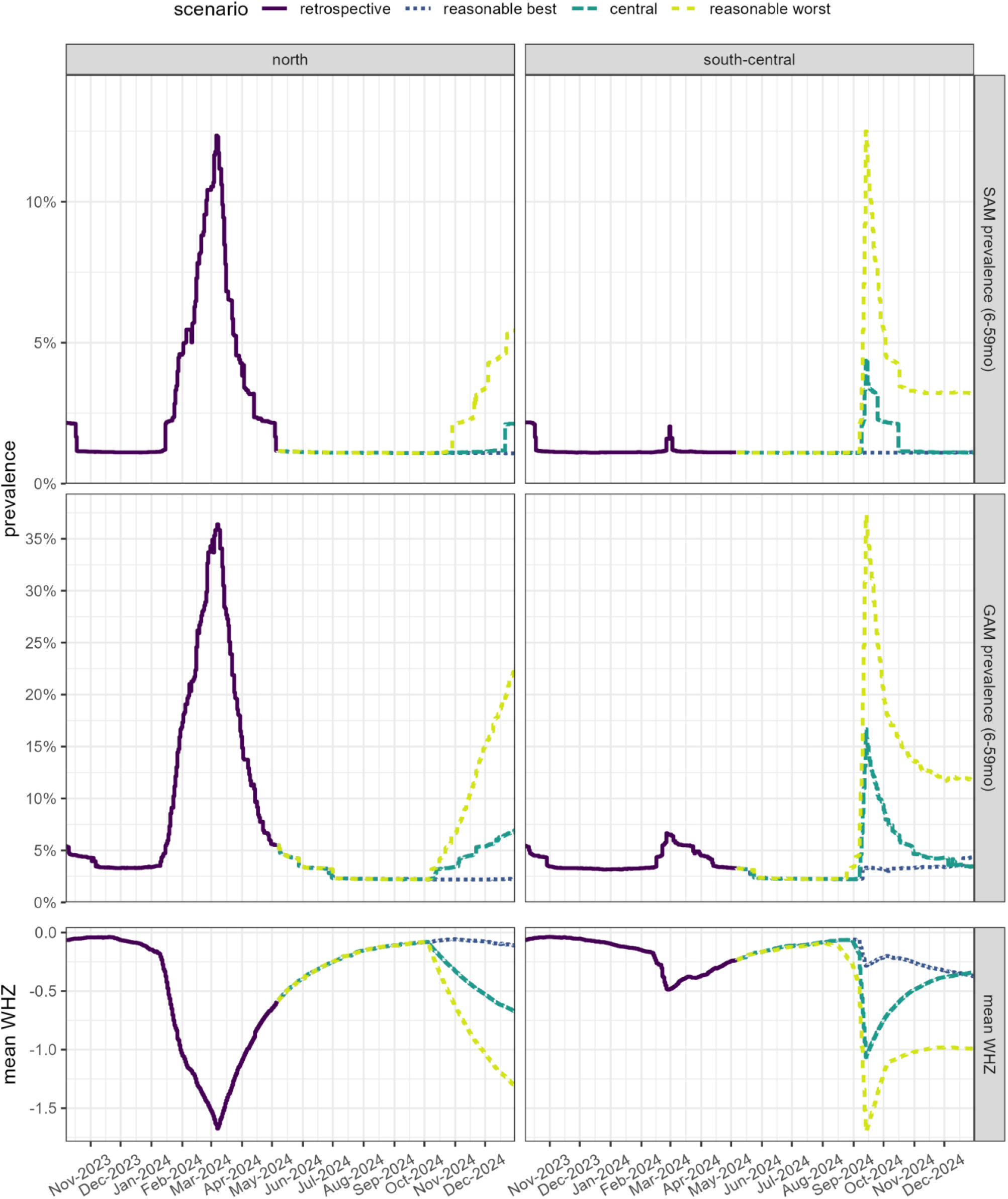
Retrospective estimates and scenario projections of SAM and GAM prevalence (children 6 to 59mo), GAM prevalence (6 to 23mo) and mean WHZ (6 to 59mo), by region and scenario, after simulating 100 cohorts of 100 children each.

## Discussion

### Main findings

To our knowledge this is the first instance of a mechanistic model that estimates expected acute malnutrition burden on the basis of caloric intake while accounting for obvious modulating factors. As such, our paper should mainly be viewed as introducing the model and reporting on its first application in a real-life crisis, thereby showcasing its current potential and limitations (see below).

The model accurately predicts the median WHO growth standard if parameterised with recommended caloric intake and, calibrated to pre-war data, reproduces expected distributions of anthropometric indices, albeit with a possible, small underestimation of variance and thus SAM/GAM prevalence. It suggests a strong role for adult caloric sacrifice as a coping strategy during periods of food scarcity.

Some 95% of households in Rafah governorate employed food security coping strategies with caretakers often skipping meals to feed their children [39].

The model suggests that in northern Gaza, food scarcity during January-March 2024 caused a rapid increase in acute malnutrition, reversed by May likely thanks to improved food availability [11]. These retrospective estimates for Gaza seem broadly in line with available ground data (Figure 5), but the latter are not a suitable dataset for validation as they do not arise from representative sample surveys and rely on MUAC-based measurement; moreover, ground data from northern Gaza during the first half of 2024 are particularly scant. Retrospective model estimates also feature wide uncertainty. Scenario projections suggest that, in both northern and south-central governorates, a substantial decline in food availability (unlike at the war’s outset, not mitigated by pre-existing stores [11]) could result in a serious nutritional emergency.

**Figure 5.**
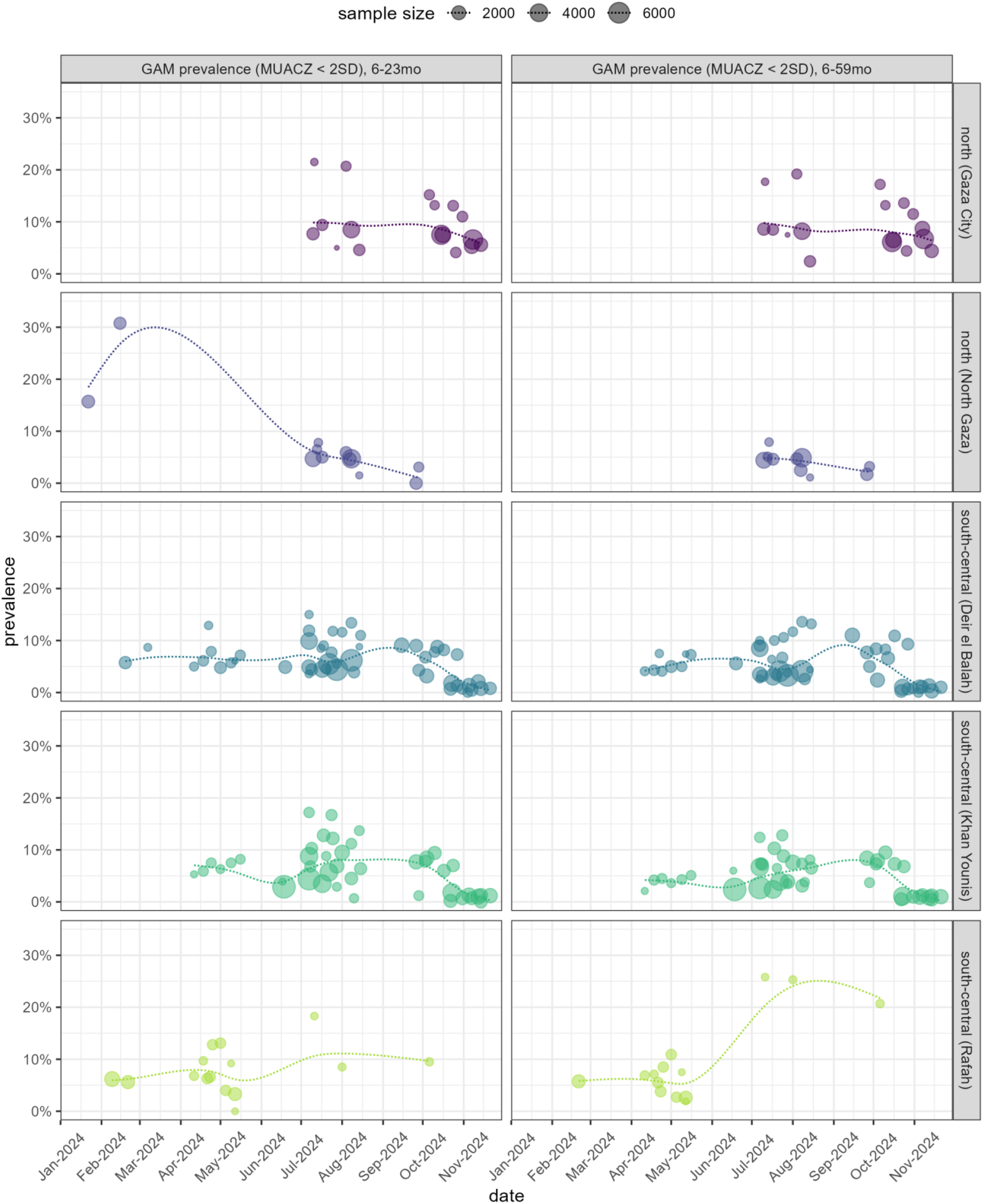
Point estimates of GAM prevalence (based on MUAC Z-score < 2SD) from ground screenings, by age group (some screenings targeted children 6 to 23mo only). Each data point is centred at the mid-point of the data collection period (usually spanning 2-3 weeks) and its size is a function of the number of children screened. To better visualise trends, we fitted a smooth spline, weighted by number screened, to periods covered by data.

### Limitations

In addition to being computationally intensive, the model has several limitations, of which the most important are emphasised here. Upstream, the model ignores food waste and access inequity; adult caloric sacrifice is modelled statically, but may in fact be function of how long adults have already been depriving themselves and/or of children’s nutritional status at the time; adding a sub-model of adult weight [52] while tracking its evolution through primary data would help improve this model component. The model’s calibration to represent inter-individual variability may not successfully replicate growth curves outside Gaza, and is unlikely to be biologically plausible, though its performance can be tested on new growth monitoring data.

Our treatment of risk factors is incomplete. We only feature half of the known vicious cycle of infection and malnutrition (infection leading to malnutrition, but not the reverse), omit epidemic and other endemic infections and the energy cost of infection through symptoms such as fever. We also ignore the higher risk of infection (especially diarrhoea) among formula-fed versus breastfed infants, e.g. due to contaminated water or bottles and the increased frequency of low birth weight, thereby probably under-estimating infant malnutrition. Lastly, in a protracted crisis our model would eventually deviate from reality because it does not predict changes in height, i.e. stunting.

The specification of model inputs for Gaza also suffers from considerable uncertainty. We made strong assumptions about caloric intake since May 2024, the extent of adult caloric sacrifice and other factors such as ARI and diarrhoea prevalence, observed values of which may have suffered from some misclassification. While the retrospective analysis is at least not grossly inconsistent with ground-observed malnutrition prevalence, this does not necessarily indicate model validity, but merely that reasonable model inputs can reconstruct a plausible trajectory of nutritional status in Gaza.

## Conclusions

An accurate model to project malnutrition in humanitarian responses on the basis of various input data, available or assumed, would complement nutritional surveillance based on primary data collection and could considerably enhance situational awareness by enabling near-real-time estimation including for populations that are hard to access and thus survey. The model’s forward-projections can potentially support decision-making by exploring the consequences of worsening food security, hostile actions such as obstruction of food delivery and humanitarian interventions to improve food access and support child nutrition. Projections can also support planning, e.g. by quantifying the expected incidence of SAM and MAM requiring treatment.

While the model presented here appears to accurately replicate both the WHO growth standards and pre-war anthropometry in Gaza, further validation of its predictions against primary data from other settings is warranted. Further model development should, at a minimum, seek to represent food inequity and a more dynamic relationship between scarcity and caloric sacrifice than featured so far; account for the synergy between infection and malnutrition; and better specify neonatal and infancy factors including low birth weight and incomplete breastfeeding. Parameter values (e.g. effect of infectious disease on intake) should be informed by more thorough literature review. Data on the evolution of adult weight during the crisis could, in particular, help to validate the adult sacrifice sub-model.

As regards Gaza, the model suggests that past periods of food scarcity resulted in sizeable increased in malnutrition prevalence. Scenario projections indicate that, during the last quarter of 2024, the population was on the brink of a serious nutritional emergency, with the balance between moderate and severe deteriorations in nutrition potentially resting on small differences in caloric availability and adults’ ability to deprive themselves so as to feed children.

## Data Availability

Data and code for implementation in R are publicly available online on https://github.com/francescochecchi/nut_cal_model_gaza

## Acknowledgments

We are grateful to UNRWA for sharing growth monitoring data and to Mija-Tesse Ververs (Center for Humanitarian Health, Johns Hopkins University) for technical advice. This research was conducted with the support of the UK Humanitarian Innovation Hub (UKHIH) and its donor, the UK Foreign, Commonwealth & Development Office (FCDO).

