## Supplementary Material for "Evolution of child acute malnutrition during war in the Gaza Strip, 2023-2024: retrospective estimates and scenario-based projections"

#### Methods

##### Growth curve estimation

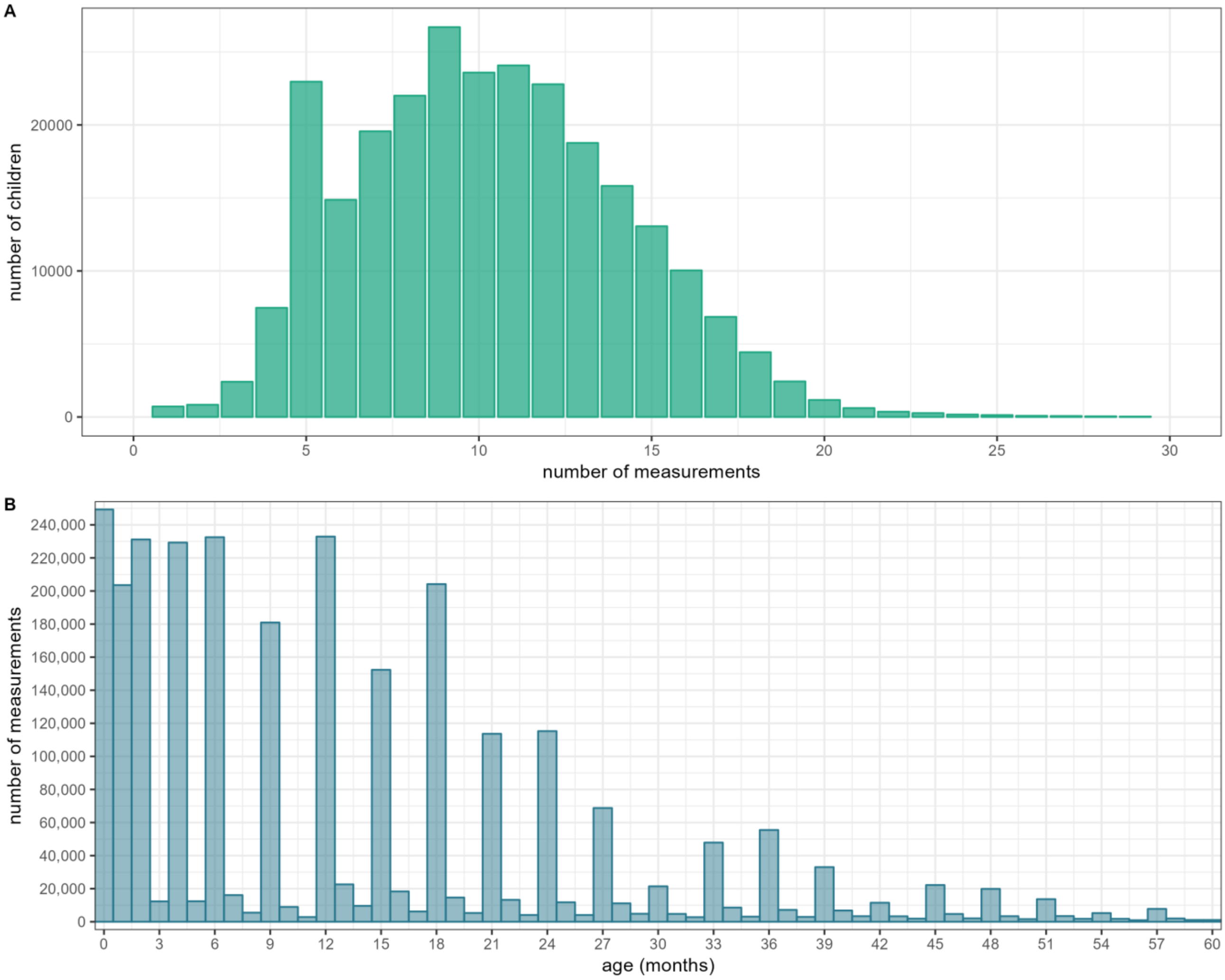


Figure S6. A: Distribution of the number of longitudinal growth monitoring measurements per child. B: Number of growth monitoring observations by age.

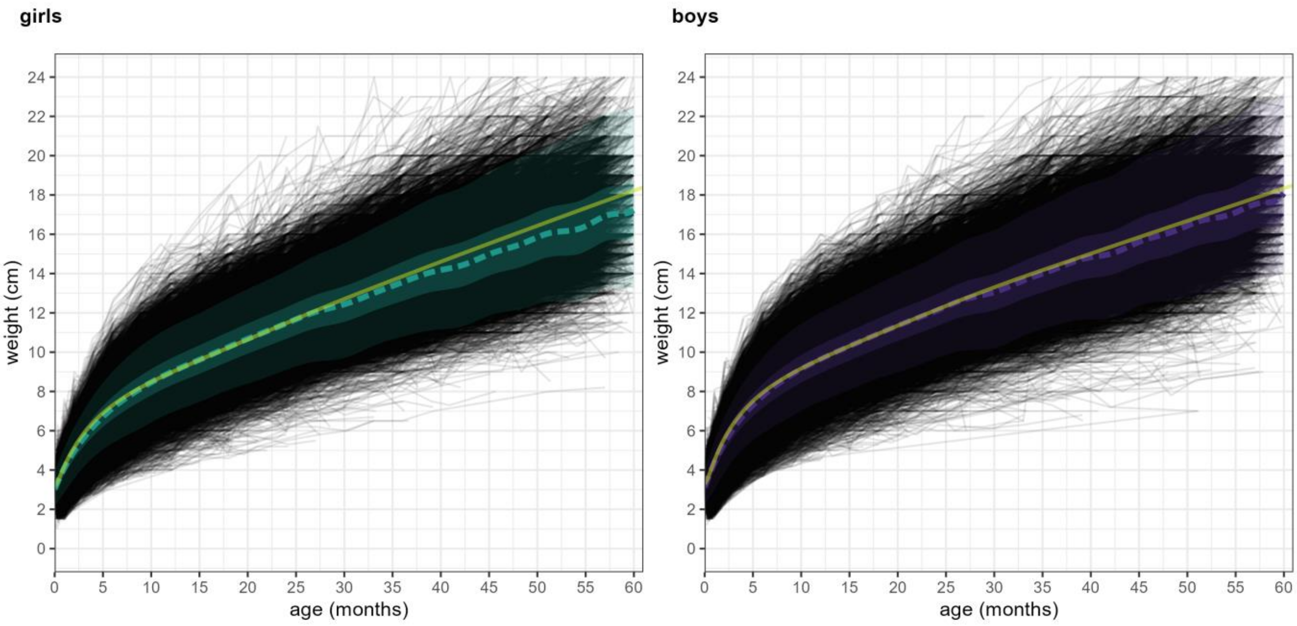


Figure S7. Weight-for-age growth curves for pre-war Gaza, by sex. Each grey segment is an individual child's trajectory. The dotted line indicates median weight-for age values in Gaza, while the solid yellow line is the median of the WHO 2006 standards for a healthy well-nourished population. Shaded areas denote quantiles of the distributions within ±1 and ±2 standard deviations, respectively.

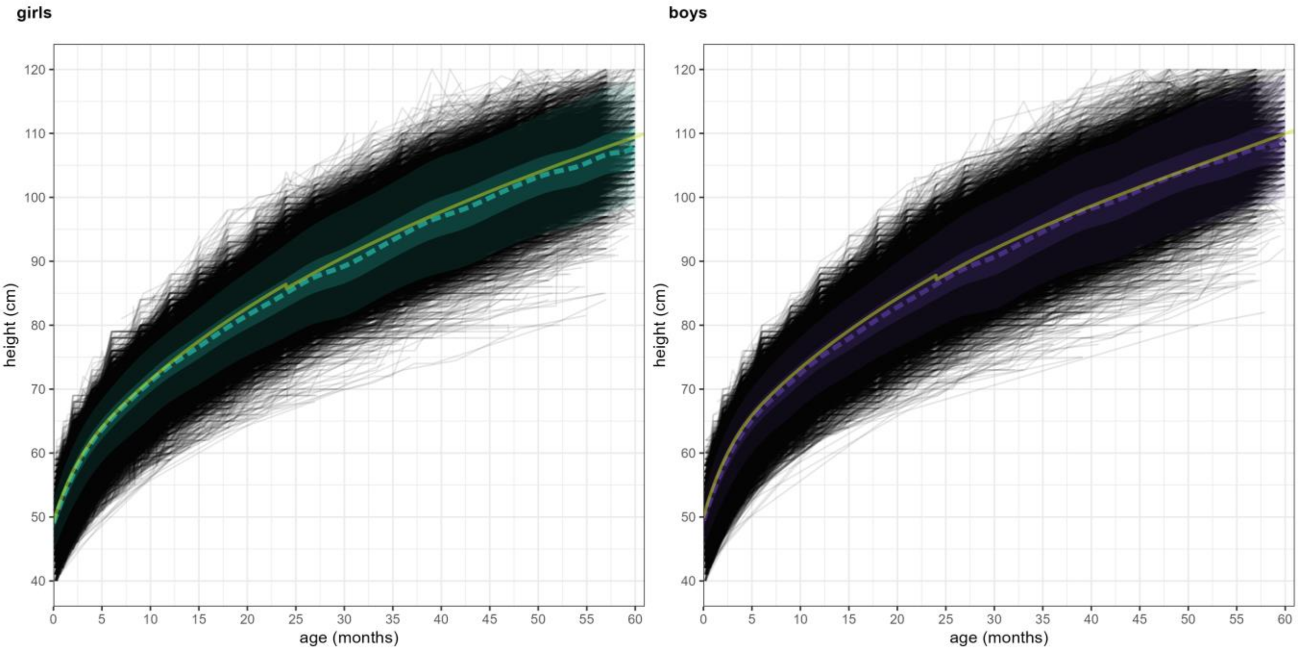


Figure S8. Height-for-age growth curves for pre-war Gaza, by sex. Each grey segment is an individual child's trajectory. The dotted line indicates median height-for age values in Gaza, while the solid yellow line is the median of the WHO 2006 standards for a healthy well-nourished population. Shaded areas denote quantiles of the distributions within ±1 and ±2 standard deviations, respectively. Note that the small disconnect around age 25mo is due to the switch from horizontal (length) to vertical (height) measurement.

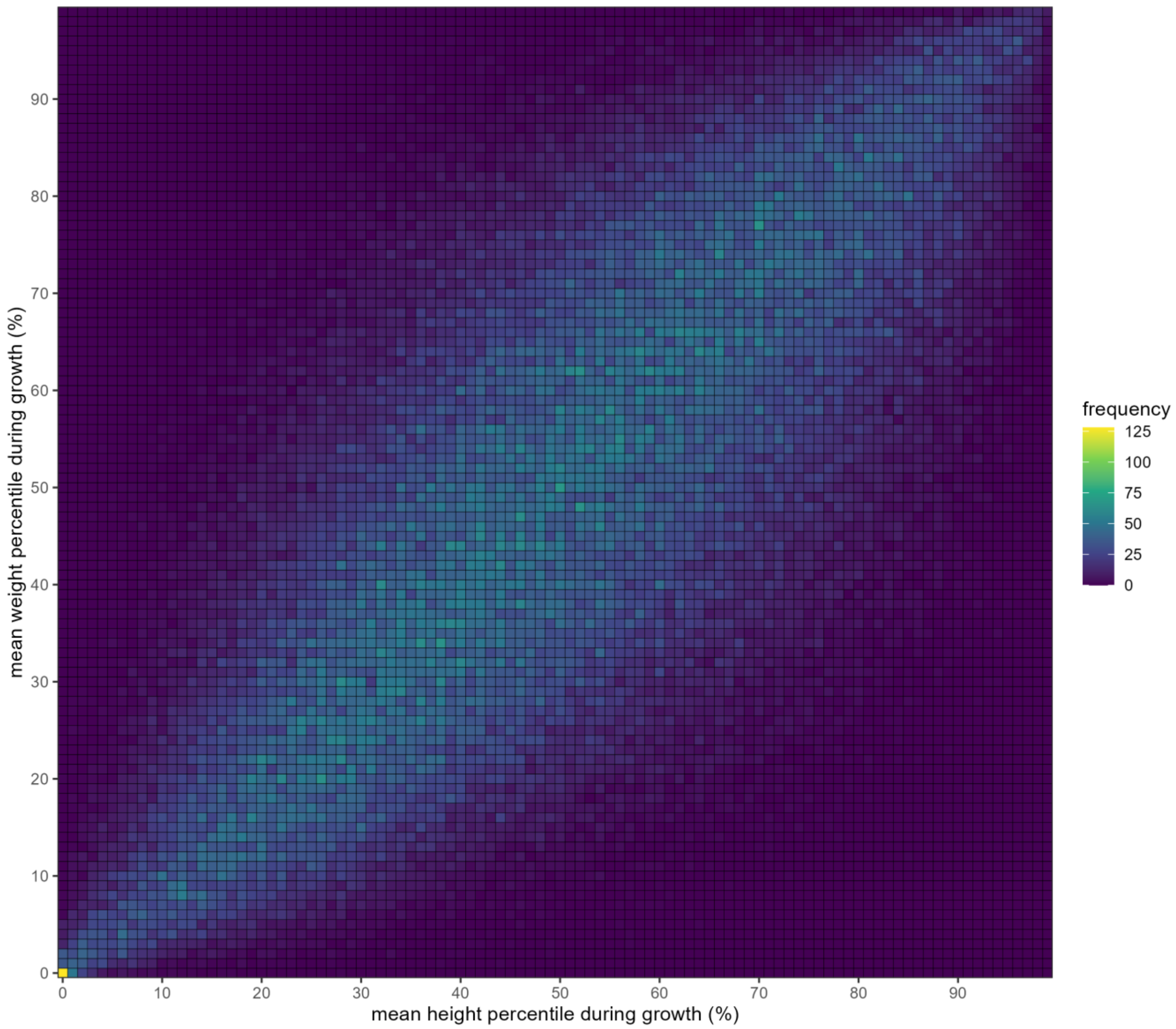


Figure S9. Observed correlation between the mean weight percentile and mean height percentile of children over their longitudinal growth monitoring observations. Each cell contains the number of children within that height-weight combination.

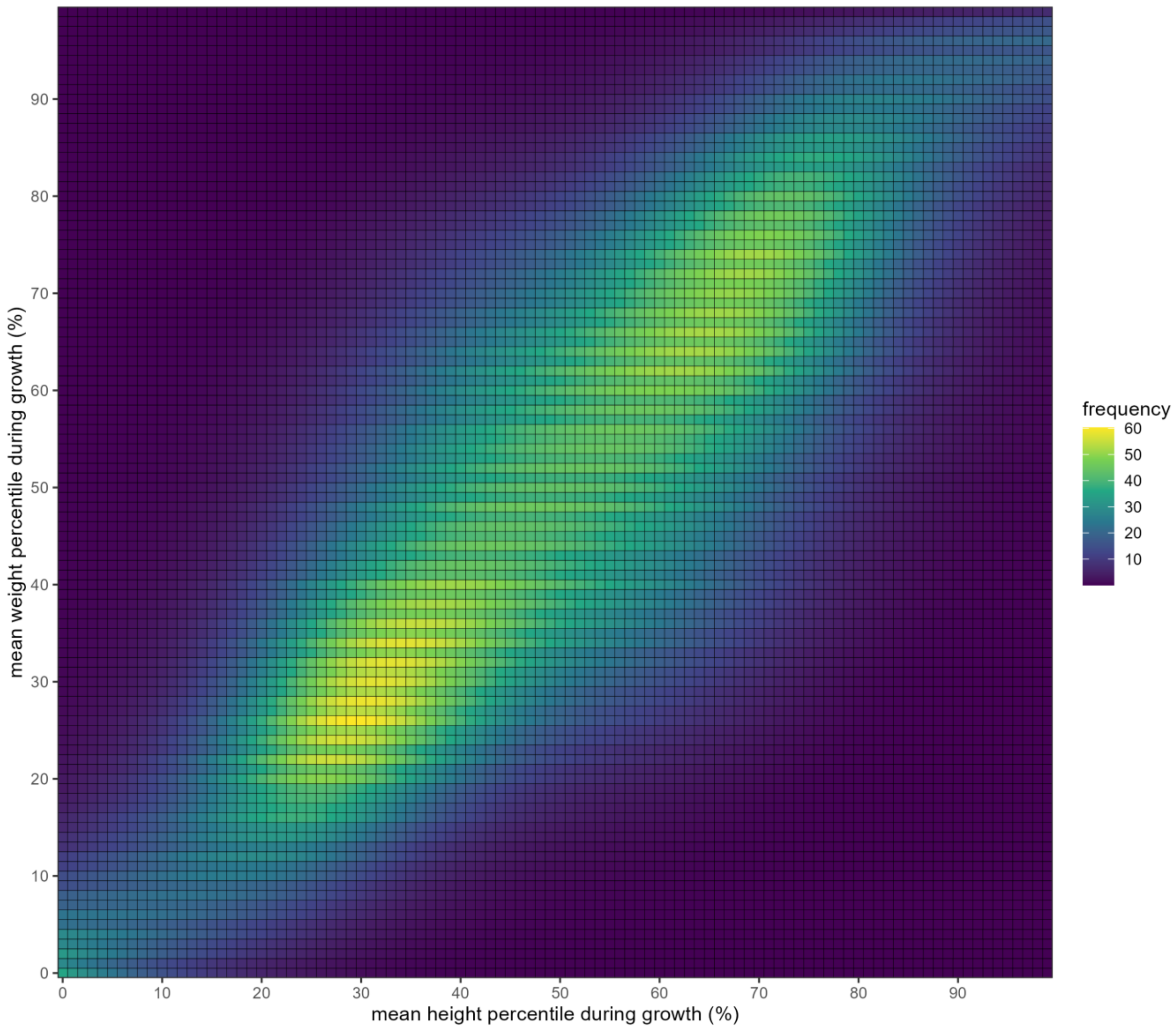


Figure S10. Fitted correlation between mean weight and height percentiles, based on a generalised additive model.

#### Weight sub-model

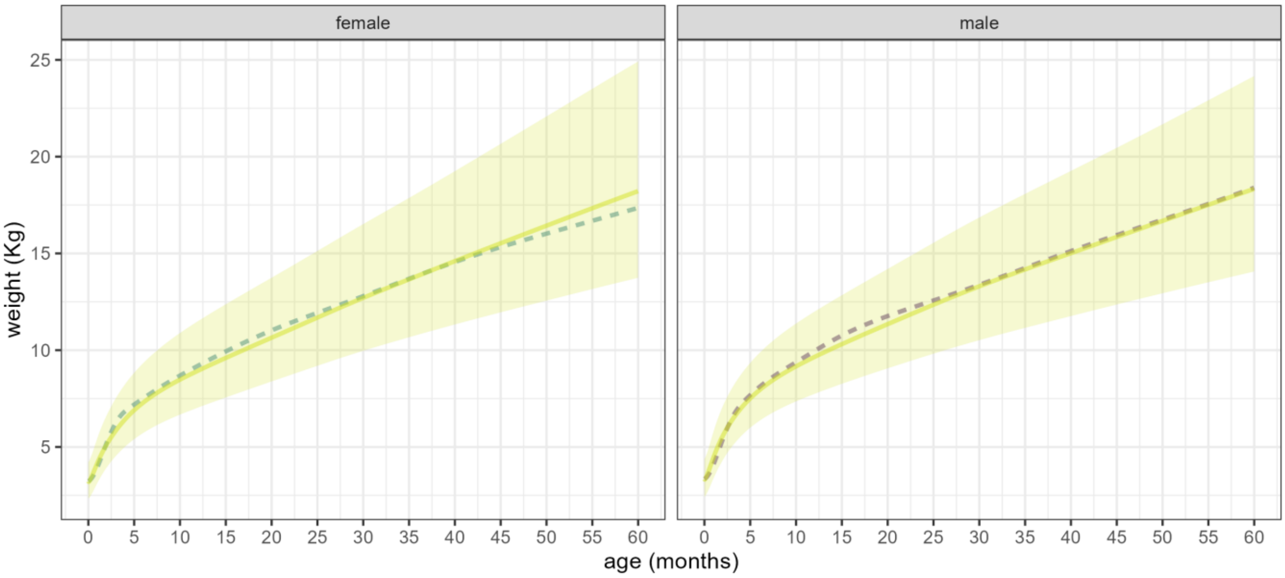


Figure S11. Weight model predictions by age and sex (dotted line), superimposed onto the mean (solid yellow line) and area within 2 standard deviations (yellow-shaded region) of the WHO growth standards.

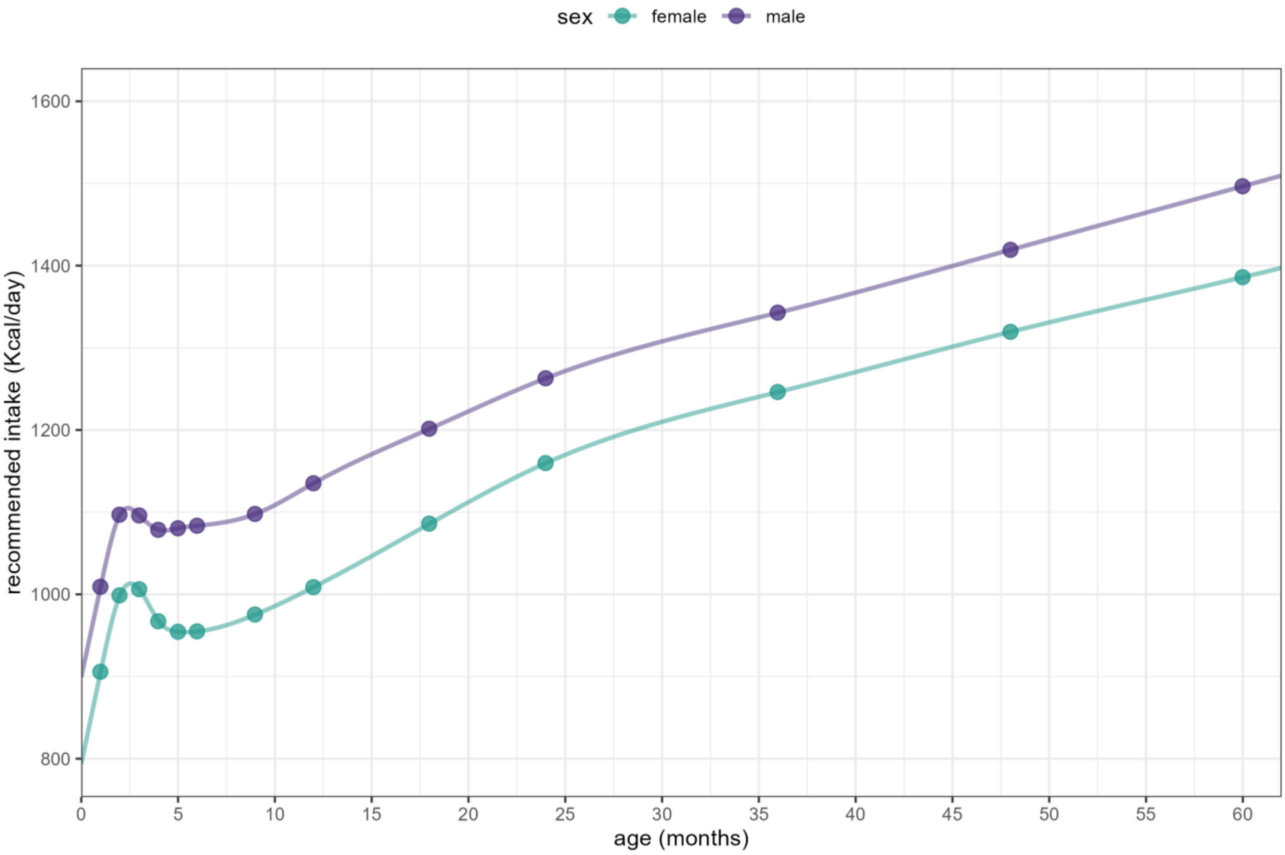


Figure S12. Model-predicted recommended intake by age and sex. Points indicate the values of age at which the model's analytic solution was applied. The interpolation line is a smoothed spline.

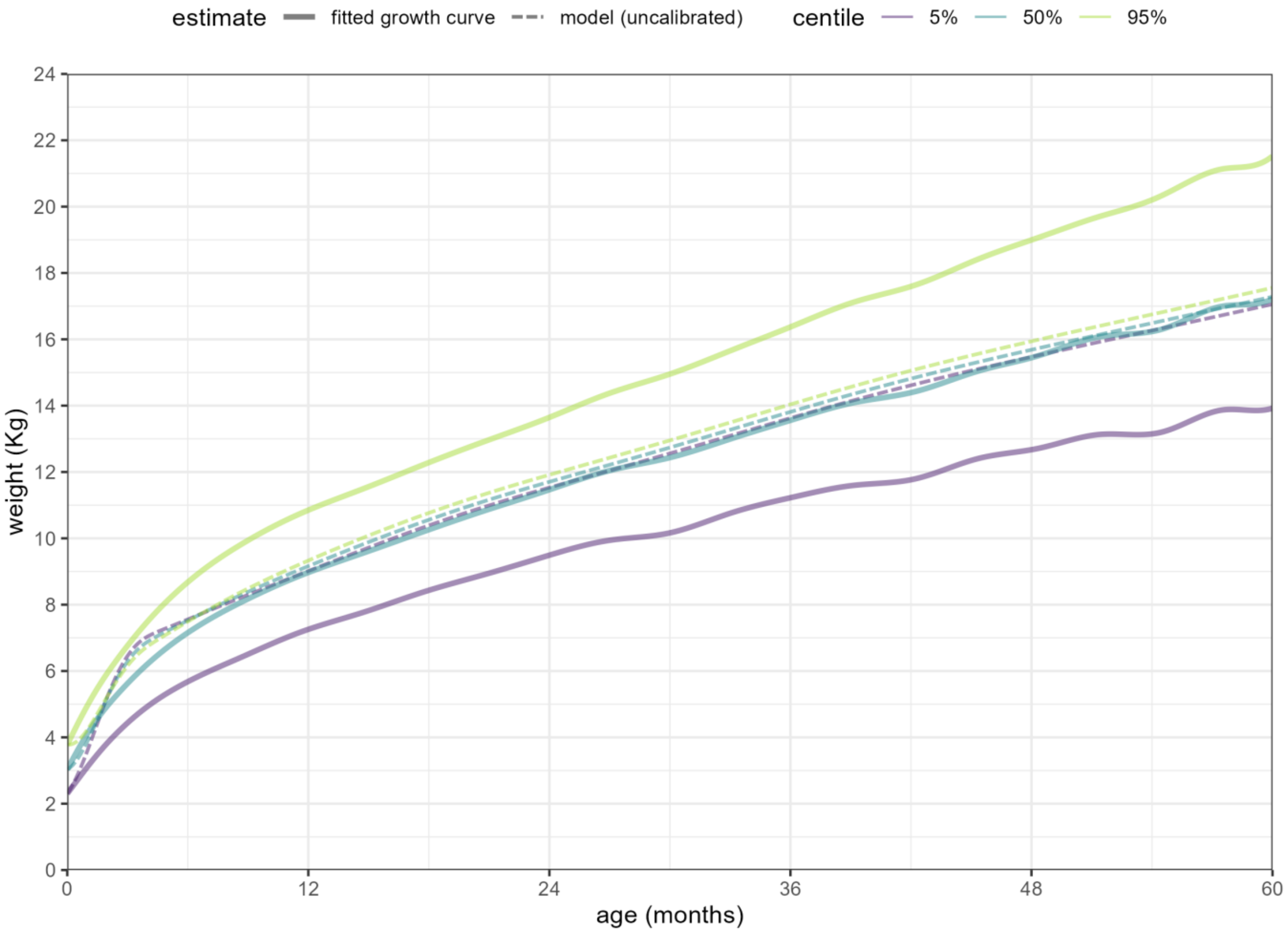


Figure S13. Uncalibrated weight model predictions (dotted lines) versus fitted growth curves for the 5th, 50th (median) and 95th weight-for-age percentile among Gazan girls before the war. As shown, even children who start out at extreme percentiles are predicted to rejoin the median very soon after birth, resulting in minimal model-predicted variance.

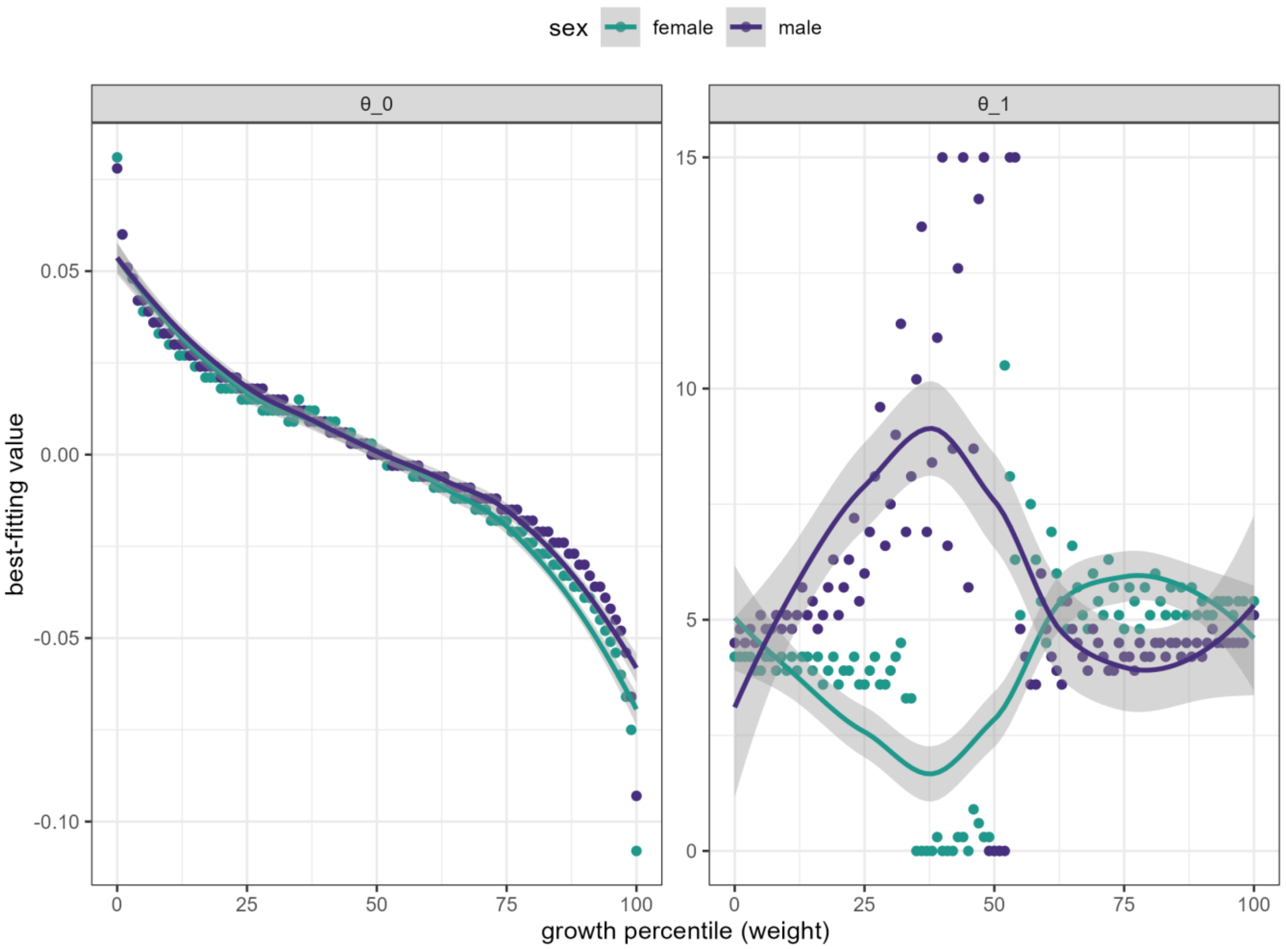


Figure S14. Maximum-likelihood (lowest root sum of squares) value of each unknown parameter (left: $\vartheta_{0,s,w}$; right: $\vartheta_{1,s,w}$), by weight percentile and sex. Each dot shown the best-fitting value, and the shaded solid line indicates a smoothed trend. Note that $\vartheta_{1,s,w}$ is uninformative at moderate percentile values, i.e. a range of values provide very similar goodness-of-fit, explaining the observed likelihood pattern.

#### Effect of infectious disease

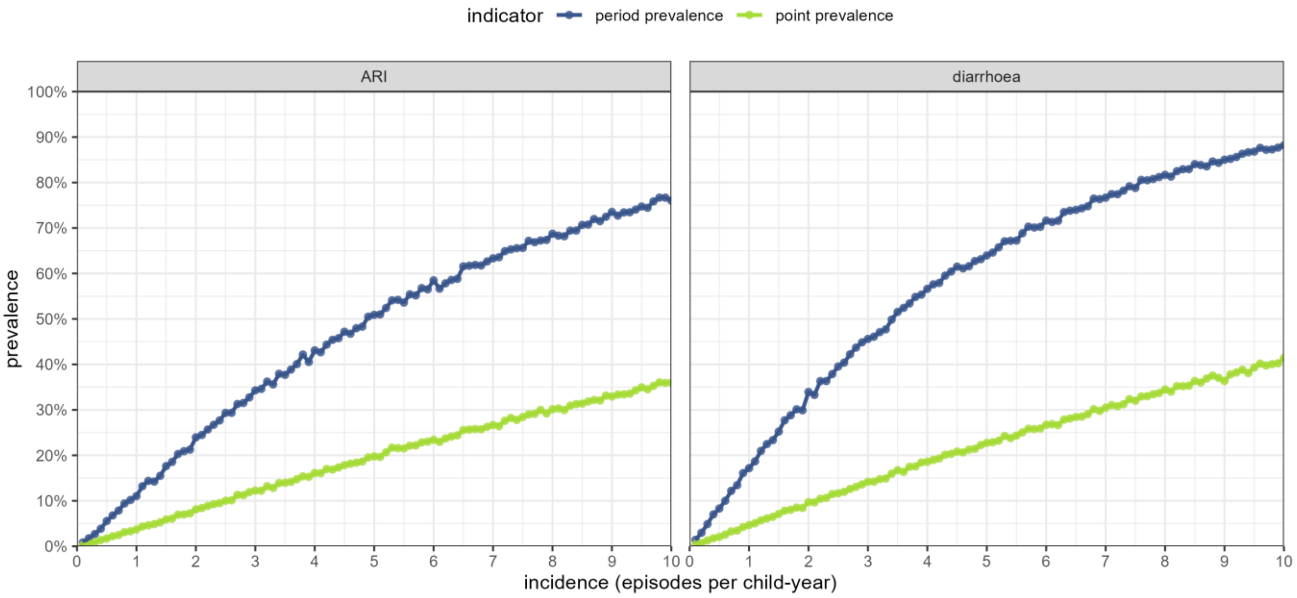


Figure S15. Estimated relationship between annual incidence, point prevalence and period prevalence over the previous two weeks, for both acute respiratory infection (left) and acute watery diarrhoea (right).

#### Effect of incomplete breastfeeding

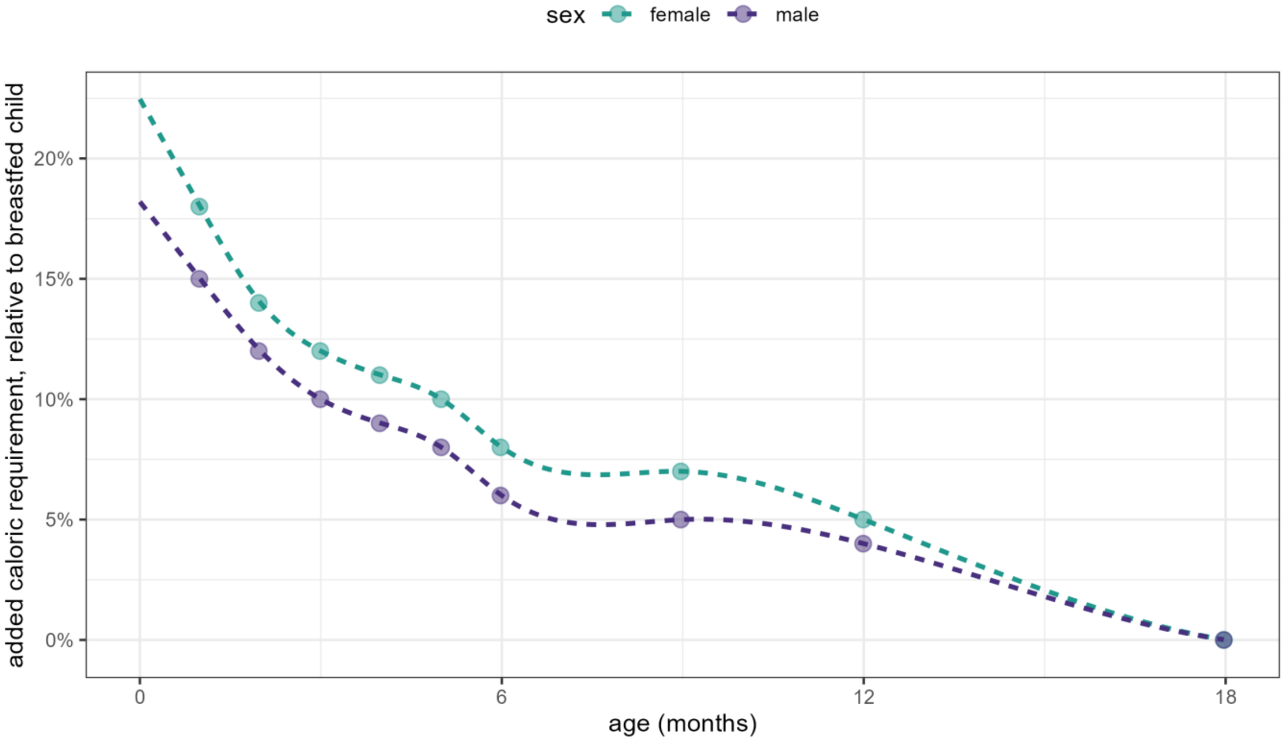


Figure S16. Percent increase in caloric requirement by age for a formula-fed child, relative to an exclusively breastfed child of the same age and sex. Data points are as provided by the Food and Agriculture Organisation; dotted lines are smoothed spline interpolations.

#### Crisis-specific data sources: pre-war intake

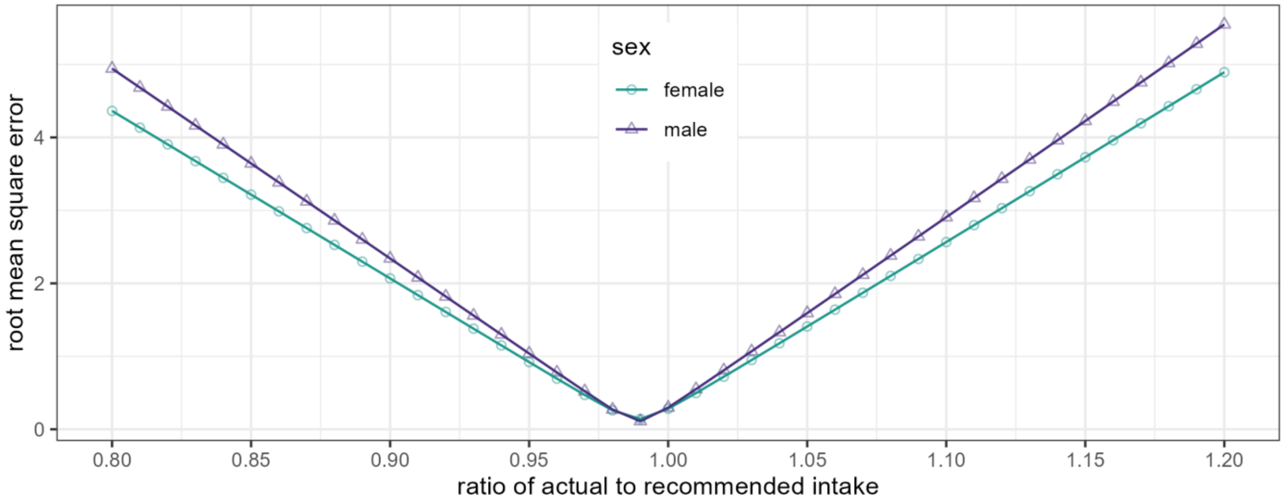


Figure S17. Root mean square error of candidate values of the ratio of actual to recommended intake (Kcal/day) among children before the war.

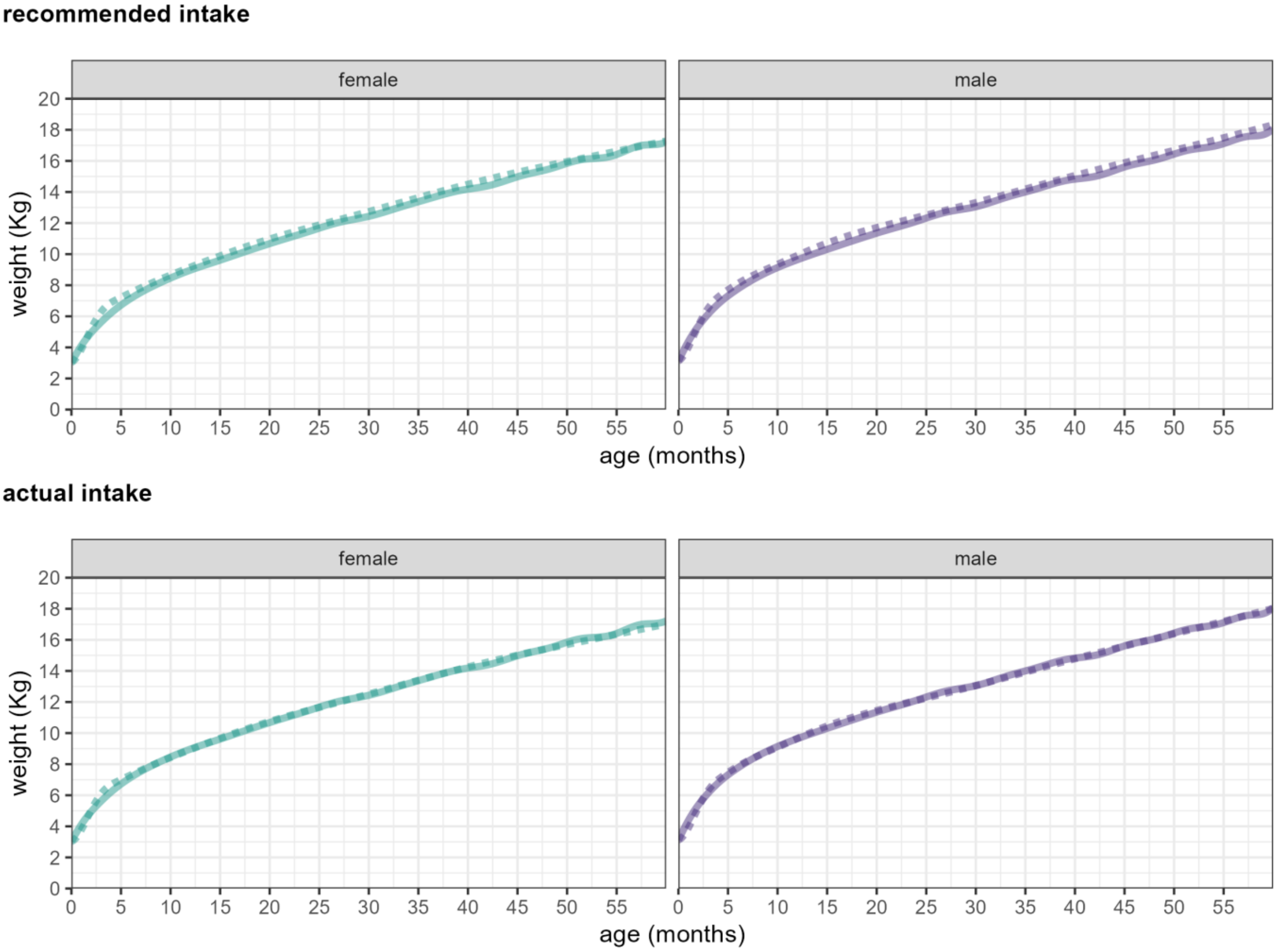


Figure S18. Weight model predictions (dotted line) versus median growth curves (solid line) if assuming the recommended intake (top) versus the maximum-likelihood value of the actual pre-war intake (bottom).

#### Crisis-specific data sources: ARI and diarrhoea period prevalence

##### Table S3. Assumed monthly values of excess ARI period prevalence, and assumptions made.

| region | month, year | observed prevalence† | assumed baseline | notes on baseline | excess prevalence | assumptions / notes |
|---|---|---|---|---|---|---|
| north | Oct 2023 | missing | 0.02 | assumed based on pre-war seasonality | 0.00 | not enough time for excess |
| | Nov 2023 | missing | 0.04 | | 0.05 | assumed |
| | Dec 2023 | missing | 0.06 | | 0.09 | |
| | Jan 2024 | 0.47 | 0.08 | MICS (Dec-Jan 2019-2020) | 0.15 | divide by two, then subtract baseline |
| | Feb 2024 | 0.31 | 0.06 | assumed based on pre-war seasonality | 0.09 | |
| | Mar 2024 | 0.33 | 0.04 | | 0.12 | |
| | Apr 2024 | 0.14 | 0.02 | | 0.05 | |
| | May 2024 | 0.05 | 0.01 | | 0.02 | |
| | Jun 2024 | 0.14 | 0.01 | | 0.06 | |
| | Jul 2024 | 0.08 | 0.01 | | 0.03 | |
| | Aug 2024 | 0.00 | 0.01 | | 0.00 | |
| | Sep 2024 | 0.00 | 0.01 | | 0.00 | |
| | Oct 2024 | missing | 0.02 | | 0.00 | assumed, as for 2023 |
| | Nov 2024 | missing | 0.04 | | 0.05 | |
| | Dec 2024 | missing | 0.06 | | 0.09 | |
| south-central | Oct 2023 | missing | 0.02 | assumed based on pre-war seasonality | 0.00 | not enough time for excess |
| | Nov 2023 | missing | 0.04 | | 0.05 | assumed |
| | Dec 2023 | missing | 0.06 | | 0.09 | |
| | Jan 2024 | 0.61 | 0.08 | MICS (Dec-Jan 2019-2020) | 0.22 | divide by two, then subtract baseline |
| | Feb 2024 | 0.46 | 0.06 | assumed based on pre-war seasonality | 0.17 | |
| | Mar 2024 | 0.29 | 0.04 | | 0.11 | |
| | Apr 2024 | 0.14 | 0.02 | | 0.05 | |
| | May 2024 | 0.08 | 0.01 | | 0.03 | |
| | Jun 2024 | 0.04 | 0.01 | | 0.01 | |
| | Jul 2024 | 0.02 | 0.01 | | 0.00 | |
| | Aug 2024 | 0.07 | 0.01 | | 0.03 | |
| | Sep 2024 | 0.00 | 0.01 | | 0.00 | |
| | Oct 2024 | missing | 0.02 | | 0.00 | assumed, as for 2023 |
| | Nov 2024 | missing | 0.04 | | 0.05 | |
| | Dec 2024 | missing | 0.06 | | 0.09 | |

† Observed monthly prevalences are the mean of Gaza City and North City for the northern region, and of remaining governorates for the south-central region.

Table S4. Assumed monthly values of excess diarrhoea period prevalence, and assumptions made.

| region | month, year | observed prevalence† | assumed baseline | notes on baseline | excess prevalence | assumptions / notes |
|---|---|---|---|---|---|---|
| north | Oct 2023 | missing | 0.16 | assumed to remain constant year-around based on pre-war patterns | 0.00 | not enough time for excess |
| | Nov 2023 | missing | 0.16 | | 0.05 | assumed |
| | Dec 2023 | missing | 0.16 | | 0.09 | |
| | Jan 2024 | 0.57 | 0.16 | MICS (Dec-Jan 2019-2020) | 0.12 | divide by two, then subtract baseline |
| | Feb 2024 | 0.35 | 0.16 | assumed to remain constant year-around based on pre-war patterns | 0.02 | |
| | Mar 2024 | 0.43 | 0.16 | | 0.06 | |
| | Apr 2024 | 0.30 | 0.16 | | 0.00 | |
| | May 2024 | 0.18 | 0.16 | | 0.00 | |
| | Jun 2024 | 0.22 | 0.16 | | 0.00 | |
| | Jul 2024 | 0.12 | 0.16 | | 0.00 | |
| | Aug 2024 | 0.17 | 0.16 | | 0.00 | |
| | Sep 2024 | 0.25 | 0.16 | | 0.00 | |
| | Oct 2024 | missing | 0.16 | | 0.00 | assumed, as for 2023 |
| | Nov 2024 | missing | 0.16 | | 0.05 | |
| | Dec 2024 | missing | 0.16 | | 0.09 | |
| south-central | Oct 2023 | missing | 0.16 | assumed to remain constant year-around based on pre-war patterns | 0.00 | not enough time for excess |
| | Nov 2023 | missing | 0.16 | | 0.03 | assumed |
| | Dec 2023 | missing | 0.16 | | 0.07 | |
| | Jan 2024 | 0.50 | 0.16 | MICS (Dec-Jan 2019-2020) | 0.09 | divide by two, then subtract baseline |
| | Feb 2024 | 0.54 | 0.16 | assumed to remain constant year-around based on pre-war patterns | 0.11 | |
| | Mar 2024 | 0.39 | 0.16 | | 0.03 | |
| | Apr 2024 | 0.30 | 0.16 | | 0.00 | |
| | May 2024 | 0.17 | 0.16 | | 0.00 | |
| | Jun 2024 | 0.13 | 0.16 | | 0.00 | |
| | Jul 2024 | 0.06 | 0.16 | | 0.00 | |
| | Aug 2024 | 0.14 | 0.16 | | 0.00 | |
| | Sep 2024 | 0.00 | 0.16 | | 0.00 | |
| | Oct 2024 | missing | 0.16 | | 0.00 | assumed, as for 2023 |
| | Nov 2024 | missing | 0.16 | | 0.03 | |
| | Dec 2024 | missing | 0.16 | | 0.07 | |

† Observed monthly prevalences are the mean of Gaza City and North City for the northern region, and of remaining governorates for the south-central region.

#### Scenario assumptions: caloric intake

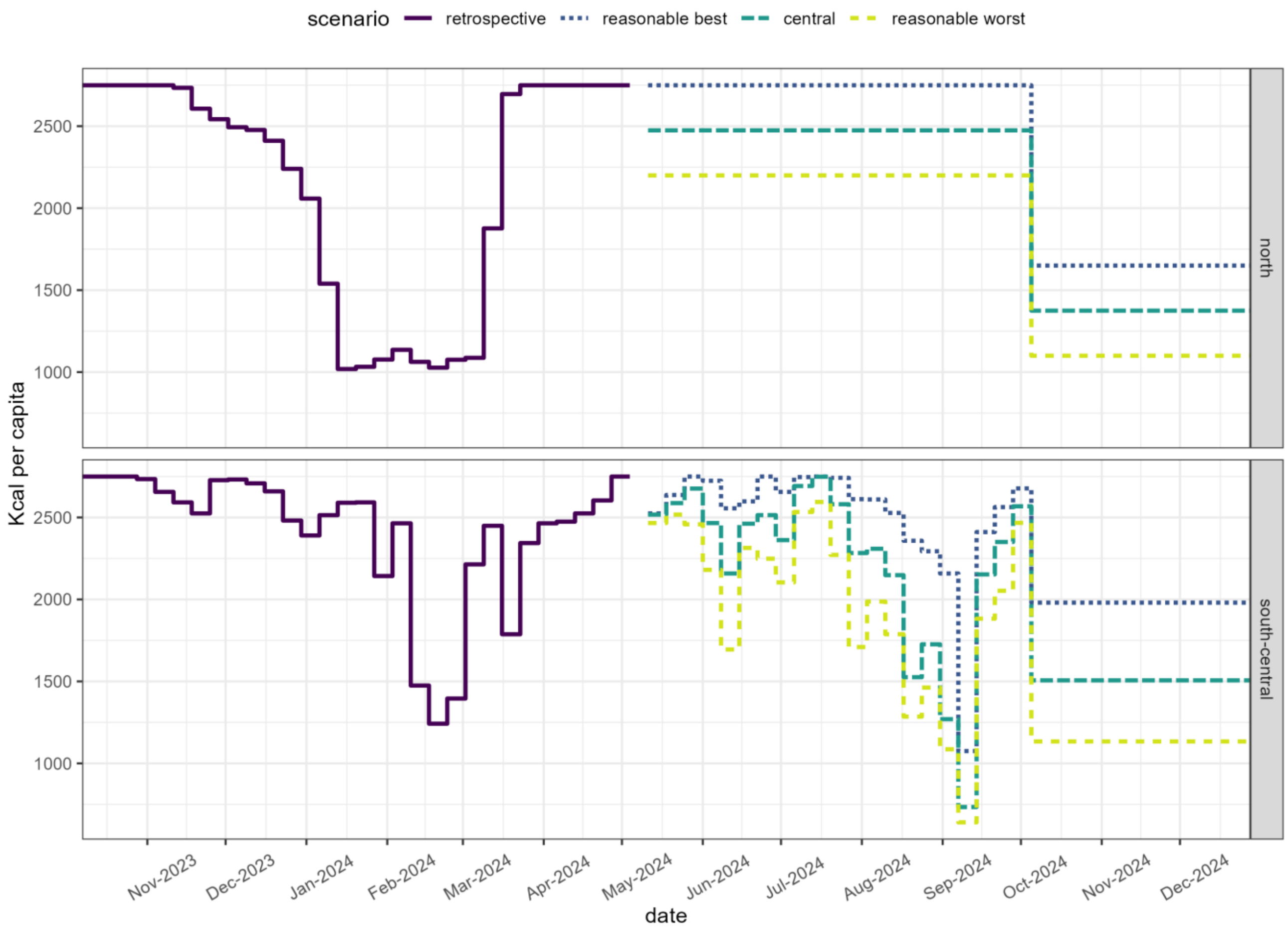


Figure S19. Retrospectively estimated and assumed values of mean caloric intake, by scenario and region.

### Results

#### Model-predicted versus observed anthropometry

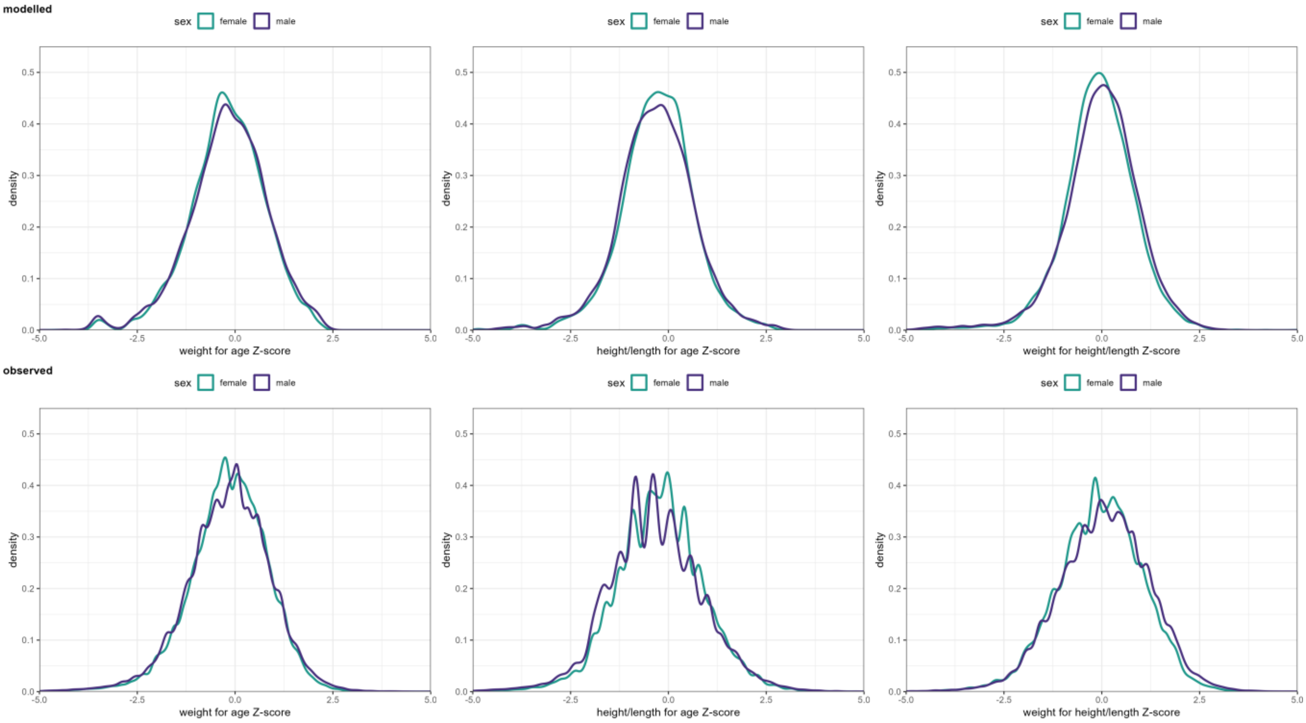


Figure S20. Comparison of pre-war model-predicted and observed (growth monitoring data) height-for-age, weight-for-age and weight-for-height Z-scores, by sex.

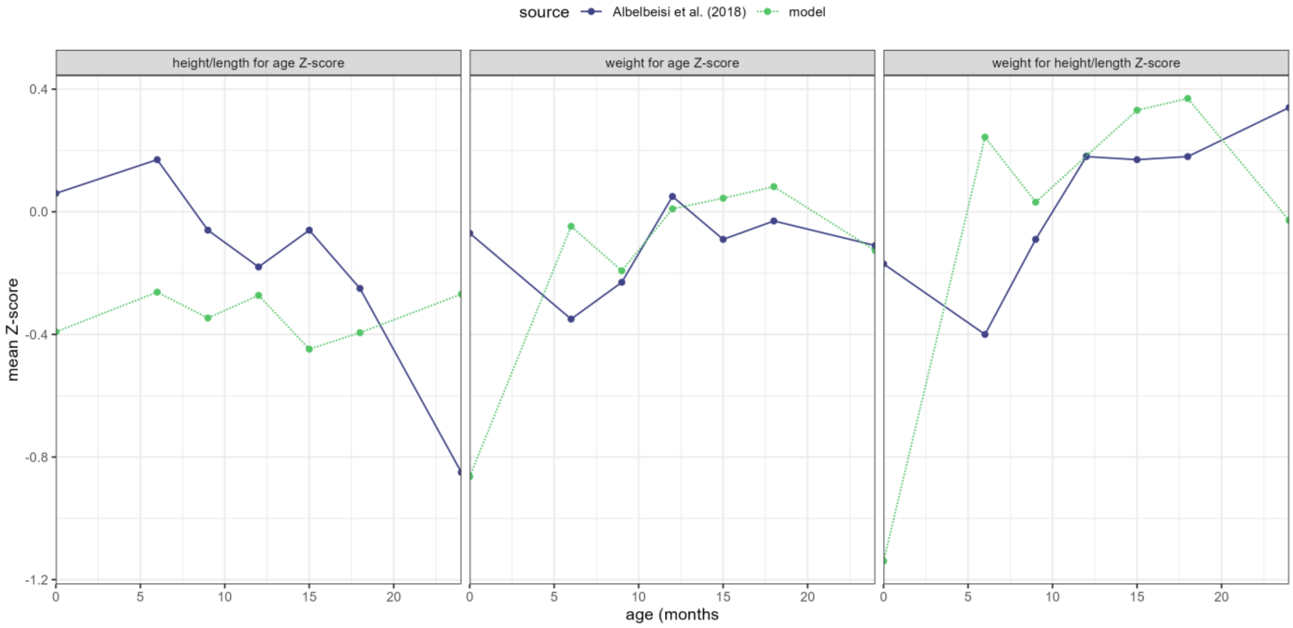


Figure S21. Pre-war mean height-for-age, weight-for-age and weight-for-height Z-scores predicted by the model and observed by Albelbeisi et al. [51] at different ages.

#### Crisis estimates

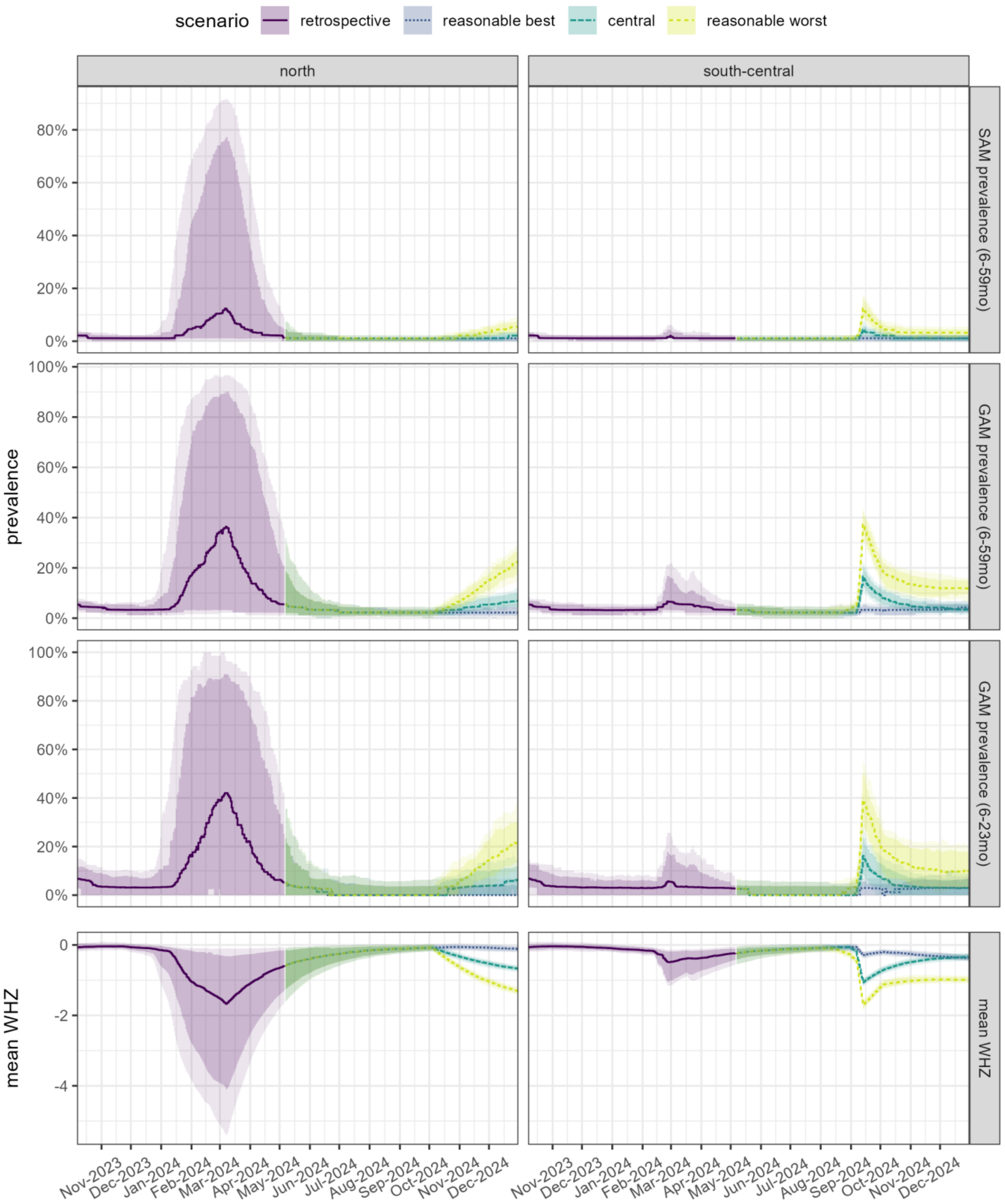


Figure S22. Retrospective estimates and scenario projections of SAM and GAM prevalence (children 6 to 59mo), GAM prevalence (6 to 23mo) and mean WHZ (6 to 59mo), by region and scenario, after simulating 100 cohorts of 100 children each. Dark-shaded areas include the 80% uncertainty interval and lighter areas the 95% uncertainty interval.
